# Adolescent pregnancy is associated with child undernutrition: systematic review and meta-analysis

**DOI:** 10.1101/2023.05.22.23290329

**Authors:** Caroline Welch, Christopher K Wong, Natasha Lelijveld, Marko Kerac, Stephanie V Wrottesley

## Abstract

Adolescent pregnancy is associated with poor foetal growth and development which increase the risk of childhood wasting and underweight. However, evidence on how young maternal age affects childhood anthropometry beyond the neonatal period is limited. This systematic review and meta-analysis examined associations between adolescent pregnancy and child wasting and underweight and explored potential underlying social and biological factors. Peer-reviewed literature published in English since 1990 was systematically searched. Eligible studies presented data on wasting and/or underweight in children (≤59 months) born to adolescent mothers (10-19, or ≤24 years where applicable) from low- and middle-income countries. Data extraction used a predefined extraction sheet. Both meta-analysis and qualitative synthesis were performed. Of 92 identified studies, 57 were included in the meta-analysis. The meta-analysis showed that children born to adolescent versus adult mothers were at a higher risk of moderate (OR: 1.12, 95% CI: 1.00-1.26 p=0.04) and severe underweight (OR: 1.21, 95% CI: 1.08-1.35 p<0.01). Associated risk of wasting was not statistically significant: (OR: 1.05, 95% CI: 0.98-1.12 p=0.17); severe wasting (OR: 1.16, 95% CI: 0.68-1.96 p=0.59). These findings were supported by the qualitative synthesis. Evidence on the potential role of biological/social factors was limited, but suggested an intermediary role of maternal nutritional status which warrants further exploration. Particularly in contexts where adolescent pregnancy remains common, interventions to both delay adolescent pregnancy and improve adolescent nutritional status could help reduce the risk of undernutrition in children and contribute to breaking the intergenerational cycle of malnutrition.

## Introduction

Approximately 11% of all births are to adolescent girls 15-19 years, of which 95% reside in low- and middle-income countries (LMICs) (Sama, Ngasa, Dzekem, & Choukem, 2017). Complications during pregnancy and childbirth are a leading cause of death for girls in this age group (World Health Organization, 2023). For those adolescent mothers who survive, the vulnerabilities faced during the postpartum period are comparatively greater than those faced by older women, and can persist through later pregnancies (James et al., 2022).

Pregnancy also restricts girls’ abilities to remain in education and increases the likelihood that they will experience unemployment, domestic violence, poverty, food insecurity and postpartum depression (Jeha, Usta, Ghulmiyyah, & Nassar, 2015; Norris et al., 2022). For adolescent mothers, such stressors can exacerbate poor physical and mental health outcomes, impacting their abilities to adequately feed and care for their children (Hentschel et al., 2022; World Health Organization, UNICEF, & World Bank Group, 2018).

Besides risks for the mother, adolescent pregnancy is associated with increased risks for the infant, including preterm delivery, low birth weight and short birth length (Fall et al., 2015; Norris et al., 2022). Evidence suggests that these risks are, at least in part, due to maternal short stature and competition for nutrients to support both adolescent and foetal growth and development during pregnancy (Norris et al., 2022). Consequences for both the mother and infant contribute to an intergenerational cycle of poverty, malnutrition and ill health (Norris et al., 2022; Ramakrishnan, Grant, Goldenberg, Zongrone, & Martorell, 2012).

Globally, an estimated 45 million children under five years of age are wasted and 85 million are underweight (Kerac et al., 2021; UNICEF, WHO, & World Bank Group, 2021), contributing to substantial burdens of mortality and morbidity in the short- and longer term (Black et al., 2013; Victora et al., 2021). Evidence supports an association between low birth weight and higher prevalence of wasting and underweight below five years of age (Aboagye et al., 2022). Experiencing an episode of wasting in early life, particularly from 0-3 months, has also been identified as a predictor of subsequent, and persistent, wasting, and concurrent wasting and stunting, as children age (Mertens et al., 2021; Mertens, Benjamin-Chung, Colford, Hubbard, et al., 2020). These associations may be further impacted by maternal nutrition and health status, socioeconomic deprivation and caregiving practices (Mertens, Benjamin-Chung, Colford, Coyle, et al., 2020).

Given the established links between adolescent pregnancy and poor foetal growth and development, and the implications of early growth faltering for childhood wasting and underweight, it can be hypothesised that being born to an adolescent mother may influence later child growth and development outcomes in LMICs. However, evidence on the impact of young maternal age on childhood anthropometry beyond the neonatal period is limited, with mixed results (Mertens, Benjamin-Chung, Colford, Coyle, et al., 2020).

This systematic review and meta-analysis aimed to examine the associations between adolescent pregnancy (10-19 years) and the risk of wasting and underweight during childhood (1-59 months), and to explore factors (biological or social) that may influence these associations, with a particular focus on the mother’s nutritional status.

## Methods

This systematic review was conducted following the process laid out by the Preferred Reporting Items for Systematic Review and Meta-analysis (PRISMA) 2009 checklist (Moher, Liberati, Tetzlaff, & Altman, 2009). The protocol was designed and approved among co-authors, and then registered with the PROSPERO International prospective register of systematic reviews in July 2022 (CRD42022327351).

### Search Strategy and eligibility criteria

The search strategy was based on the following concepts: (1) child wasting and underweight (1-59 months); (2) pregnant adolescents or adolescent mother (10-19 years), including adolescent’s nutritional status; (3) low- and middle-income countries. A detailed outline of the search strategy and terms is provided in **Supplementary Appendix 1**.

Child wasting was defined as weight-for-height z-score (WHZ) <-2, weight-for-length z-score (WLZ) <-2, mid-upper arm circumference (MUAC) <125mm, and/or bilateral oedema, and underweight was defined as weight-for-age z-score (WAZ) <-2. Only studies that disaggregated child outcomes by maternal age were eligible. Peer-reviewed literature published in English between 1990 and 21 July 2022 was included.

Due to the limited number of studies that included only children from one month of age and/or defined the youngest maternal age category as <20 years, flexibility was given to include children below one month and mothers 20-24 years, provided other eligibility criteria were met.

Studies were identified across five databases on 21 July 2022: Medline, EMBASE, Cumulative Index to Nursing and Allied Health Literature (CINAHL), Global Health and Cochrane library. All identified records were imported into EndNote (Version 20, Clarivate) and duplicates were removed. Titles and abstracts were initially screened to identify and remove studies outside the scope of review. A detailed review of full texts was then undertaken for all remaining results, against the inclusion and exclusion criteria. Studies were independently screened by two researchers. In cases where inclusion/exclusion of articles was unclear, these were discussed between co-authors.

### Data extraction

The following data were extracted from the identified studies: author(s), publication date, publication type, study design, setting, population details, outcome(s) measured, analysis method, exposure and comparator details, sample size for exposure / comparators (where provided), number of wasted and/or underweight for exposure / comparators (where provided), results exploring the potential role of other factors in the association between maternal age and wasting and/or underweight in offspring (where provided), response rate (where appropriate to study design), number of follow-ups (where appropriate to study design), follow-up rate (where appropriate to study design), intervention results (where appropriate to study design).

### Risk of bias and quality assessment

The Study Quality assessment Tools developed by the National Heart, Lung and Blood Institute (NHLBI) (National Heart Lung and Blood Institute, 2013) were used to assess the quality and risk of bias in the included studies, according to their study designs. Studies were then graded on their overall profile (good, fair, poor), with ratings being downgraded due to serious methodological issues.

### Analysis

The analysis was performed in two stages, a meta-analysis, and a qualitative synthesis.

### Meta-analysis

The meta-analysis was conducted according to the analysis plan (A1-10) presented in **Table 1**. The primary analysis (A1-4) included all studies that presented data on the prevalence of wasting and/or underweight in children 1–59 months disaggregated by maternal age. In cases where studies used the same data sources and there was clear homogeneity in the analysis methods and findings, one study was randomly selected for inclusion in the meta-analysis. Any studies excluded from the meta-analysis were still included in the qualitative synthesis.

**Table 1.** Meta-analysis plan.

| Reference | Outcome | Details |
| --- | --- | --- |
| Primary analysis |  |  |
| A1 | Moderate wasting | Includes all studies that report this outcome disaggregated by maternal age |
| A2 | Severe wasting |  |
| A3 | Moderate underweight |  |
| A4 | Severe underweight |  |
| Stratified analysis |  |  |
| A5 | Moderate and/or severe wasting <sup>a</sup> | Stratified by region |
| A6 | Moderate and/or severe underweight <sup>a</sup> | Stratified by region |
| Sensitivity analysis |  |  |
| A7 | Moderate wasting | Studies that use <20 years as the youngest maternal age / exposure cut-off |
| A8 | Severe wasting |  |
| A9 | Moderate underweight |  |
| A10 | Severe underweight |  |

Definitions: moderate wasting, weight-for-height z-score (WHZ) or weight-for-length z-score (WLZ) <-2 to -3 or mid-upper arm circumference (MUAC) 125mm to >115mm; severe wasting, WHZ or WLZ <-3 or MUAC ≤115mm; moderate underweight, weight-for-age z-score (WAZ) <-2 to -3; severe underweight, WAZ <-3
<sup>a</sup>Stratified analyses were conducted for outcomes with sufficient data available per region

Random effects meta-analyses were performed in STATA/SE 17.0 (StataCorp. 2021. *Stata Statistical Software: Release 17*. College Station, TX: StataCorp LLC) using the ‘*meta esize’* command, producing pooled estimates for the odds of wasting and underweight in offspring born to adolescent mothers versus adult mothers. Where data were disaggregated by multiple ‘adult’ maternal age categories the findings were combined to create one adult category for comparison. Pooled effects were presented as odds ratios (OR) with 95% confidence intervals (95% CI) and p-values (p). Egger tests for each pooled model were performed in STATA to assess for publication or small study bias (the null hypothesis is no bias).

Stratified analysis was conducted by region for any outcomes with sufficient data (A5-6). While stratification by children’s age category (<6 months, 6-<24 months, and 24–59 months) was also planned to identify any differences in risk of wasting and/or underweight according to life stage, this was not conducted due to a lack of appropriately age-disaggregated data.

Due to heterogeneity in the objectives of included studies, and in how maternal age was disaggregated, a sensitivity analysis was conducted to assess any differences in effects produced by the main analysis (including all studies) and a sub-analysis of studies in which the youngest maternal age category included adolescents <20 years of age (A7-10).

### Qualitative synthesis

The qualitative synthesis included all studies that met the inclusion criteria but did not present the data in a directly comparable way, e.g., they only presented adjusted ORs, or other measures of effect, or did not include a comparison group. Individual studies were assessed and summarised according to whether findings indicated an association between adolescent pregnancy (versus adult pregnancy) and childhood wasting and/or underweight. Where more than one effect estimate was reported per outcome category e.g., across models adjusted for different variables, or presenting annual trends, the final adjusted models or the most recent findings respectively were included. Handsearching of the discussion section of the final papers was undertaken to identify potential biological and/or social factors that influenced the associations, with a particular focus on those that included data on the mother’s nutritional status.

## Results

### Study selection

After deduplication. 19,368 unique references were identified (**Figure 1**). Initial title/abstract screening resulted in 694 full text articles being reviewed. In all, 602 references were excluded, primarily due to a lack of disaggregation of child outcomes (wasting and/or underweight) by maternal age. A total of 92 studies met the inclusion criteria, 57 of which presented extractable data for the meta-analysis.

**Figure 1.**
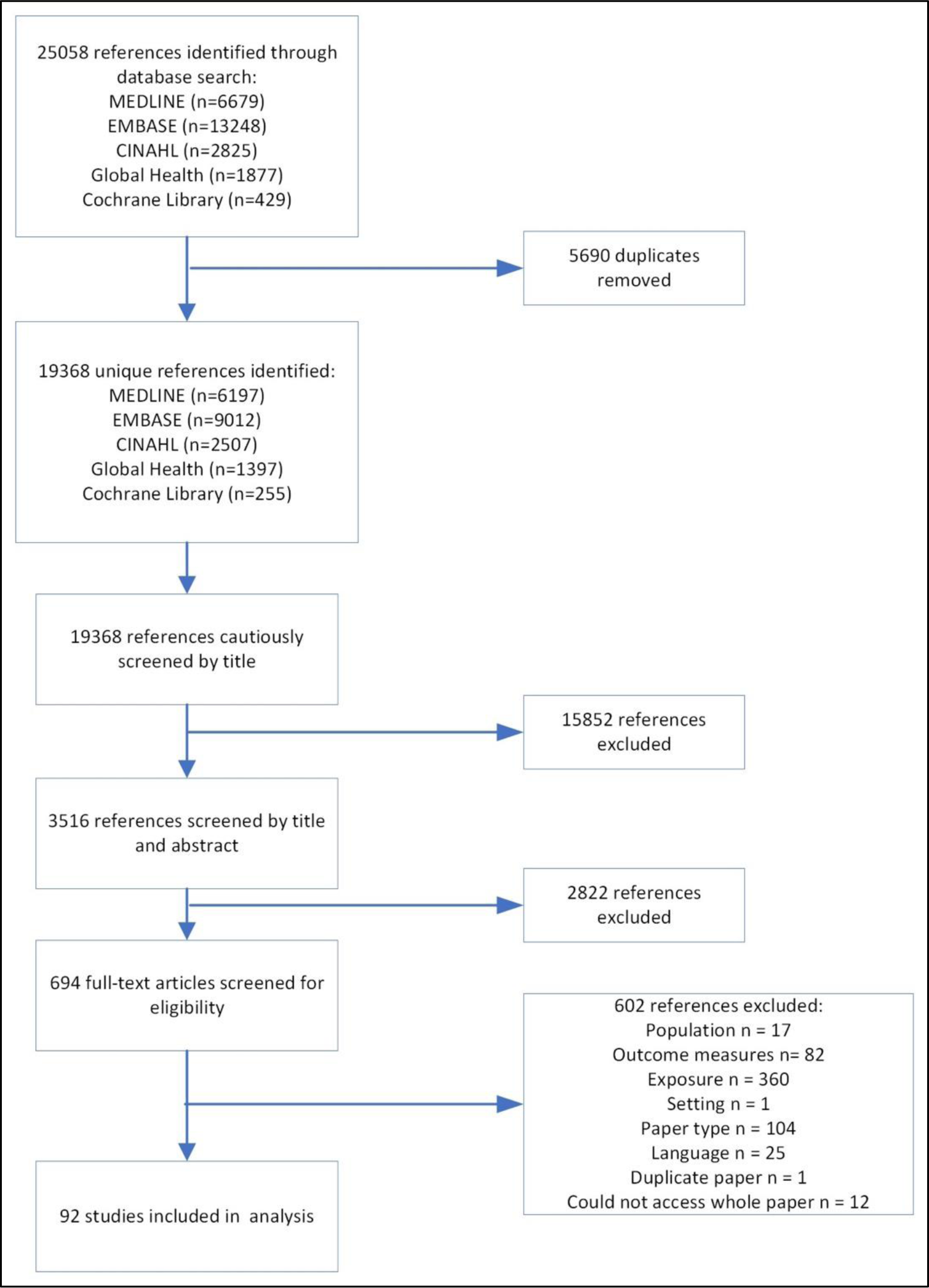
Study flow diagram.

### Study characteristics

All included studies were published between 2003 and 2022. A summary of study characteristics is provided in **Table 2**, with additional detail and full references provided in **Supplementary Appendices 2-3.** Most studies were cross-sectional in design (81/92), five were cohort studies, five were case control, and one was a randomised control trial (RCT). Most studies were conducted in Asia (50/92) and sub-Saharan Africa (SSA) (34/92). Of the remaining eight, two were conducted in South America, one in the Middle East, one in Eastern Europe, and four presented data across multiple regions.

**Table 2.** Summary of study characteristics (n=92)

|  |  |
| --- | --- |
| **Study type** |  |
| Cross-sectional | 81 |
| Cohort | 5 |
| Case control | 5 |
| Randomised controlled trial | 1 |
| **Exposure (maternal age) youngest group disaggregation** |  |
| <20 years | 69 |
| ≤24 years | 23 |
| **Outcomes reported** |  |
| Moderate wasting (WHZ or WLZ <-2 to -3 or MUAC 125mm to >115mm) | 68 |
| Severe wasting (WHZ or WLZ <-3 or MUAC ≤115mm) | 9 |
| Moderate underweight (WAZ <-2 to -3) | 71 |
| Severe underweight (WAZ <-3) | 6 |
| **Geographical location** |  |
| Sub-Saharan Africa | 34 |
| Asia | 50 |
| South America | 2 |
| Middle East | 1 |
| Eastern Europe | 1 |
| Multiple countries / continents | 4 |
| **TOTAL papers identified** | **92** |

Abbreviations: MUAC, mid-upper arm circumference; WAZ, weight-for-age z-score; WHZ, weight-for-height z-score; WLZ, weight-for-length z-score

The exposure (maternal age) was categorised in several different ways e.g., age at first birth, age at marriage and maternal age. There were also differences in disaggregation of maternal age across studies: 69 studies categorised adolescence according to the WHO definition of 10-19 years, with at least one maternal age category <20 years. The remaining studies (23) defined the youngest maternal age category as ≤24 years.

Studies reported one or more of the following outcomes: moderate wasting (WHZ <-2, WLZ <-2 or MUAC ≤125mm) (67/92), severe wasting (WHZ <-3, WLZ <-3 or MUAC ≤115mm) (9/92), moderate underweight (WAZ <-2) (72/92) and severe underweight (WAZ <-3) (6/92). No studies reported wasting assessed via oedema. One longitudinal study presented repeated anthropometric measurements (WAZ <-2, WHZ <-2) from the same cohort at 3, 6, 9, 12 and 24 months (Le Roux et al., 2019). Data for each time point have been treated as individual estimates in the analysis.

Eighteen studies examined the potential influence of other factors on observed associations between maternal age and child anthropometric outcomes: 2/18 explored biological factors, 8/18 explored social factors and 8/18 explored a combination of both. Seven studies presented data on maternal nutritional status (assessed via height, weight, body mass index (BMI), anaemia prevalence and dietary intake), disaggregated by maternal age. Three studies assessed the double burden of malnutrition (DBM) in mother-child pairs (overweight or obese mother and wasted or underweight child).

### Risk of bias and quality assessment

Of the 92 studies included, 15 were rated as ‘fair’, and the remainder as ‘good’ according to the NHLBI tools (**Supplementary Appendix 4**). Reasons for downgrading studies included a lack of adjustment for potential confounding variables and justification of sample size, as well as potential bias in the sampling process (primary studies). Given that all studies were rated as of good/fair quality, none were excluded from the analysis.

### Meta-analysis

#### Pooled analysis for child wasting

The meta-analysis included 41 studies (44 individual datapoints) with data for moderate wasting (**Figure 2**) and nine with data for severe wasting (**Figure 3**). The pooled results for all included studies showed no statistically significant association between adolescent pregnancy (≤24 years) and moderate (OR: 1.05; 95% CI 0.98-1.12 p=0.17) or severe wasting (OR: 1.16; 95% CI: 0.68–1.96 p=0.59) in children 1-59 months compared to adult pregnancy.

**Figure 2.**
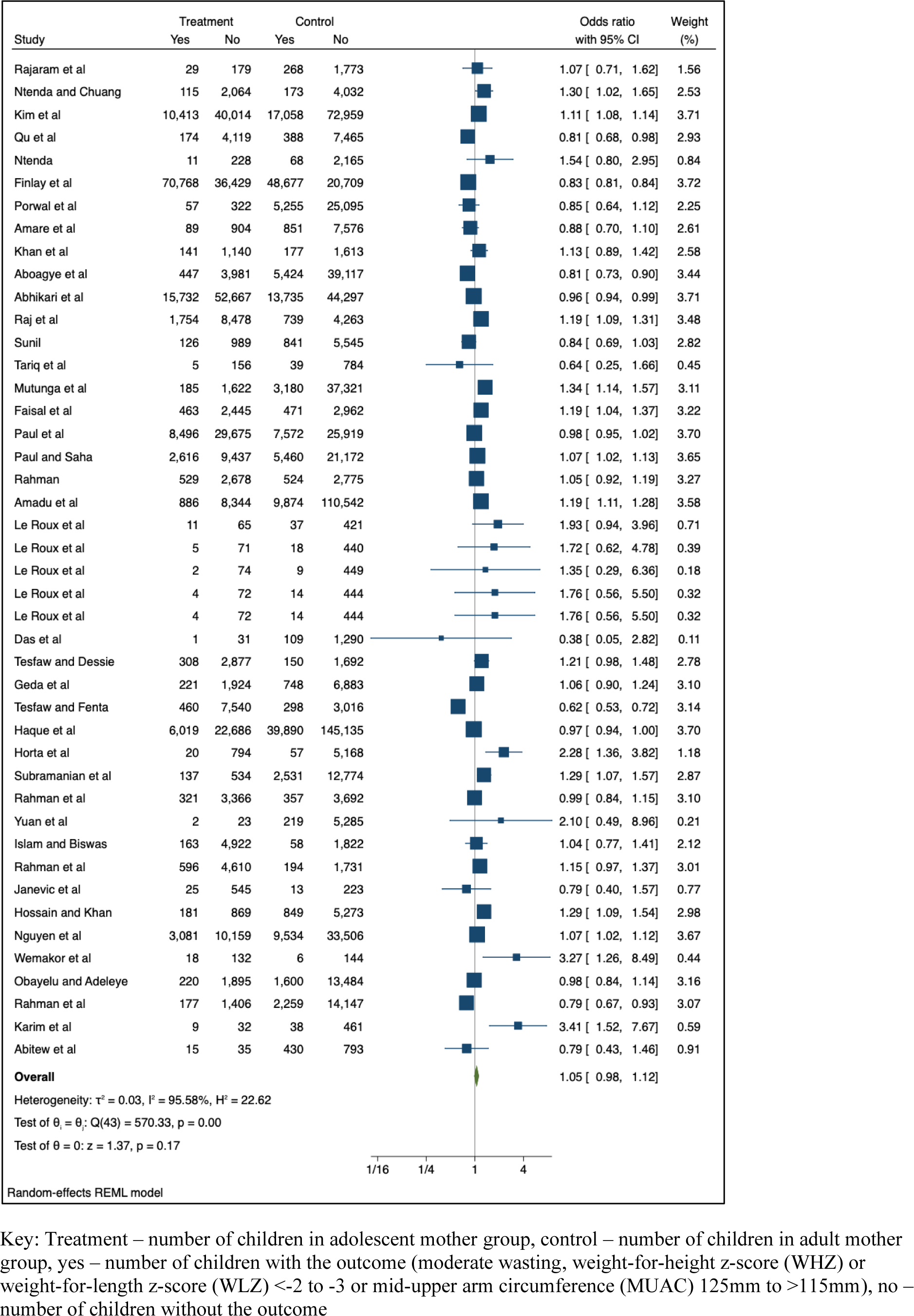
Association between adolescent pregnancy (10-24 years) and the pooled odds of moderate childhood wasting (1-59 months), versus adult pregnancy.

**Figure 3.**
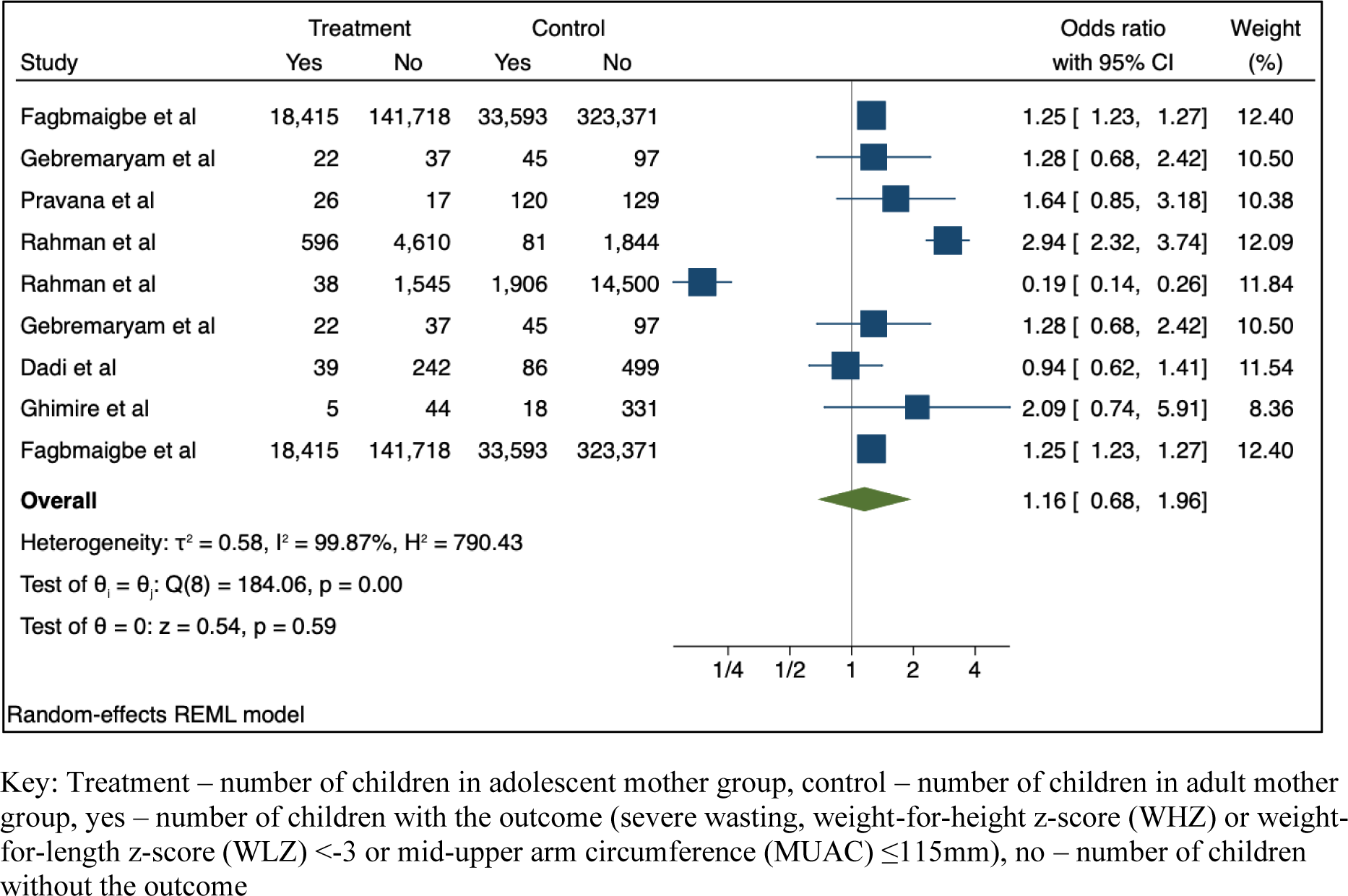
Association between adolescent pregnancy (10-24 years) and the pooled odds of severe childhood wasting (1-59 months), versus adult pregnancy.

The sensitivity analysis removed seven studies from the random effects model for moderate wasting (**Supplementary Figure 1**) and five from the model for severe wasting (**Supplementary Figure 2**). While the pooled OR indicated associations between adolescent (10-19 years) versus adult pregnancy and moderate and severe wasting, findings were not statistically significant (moderate wasting, OR: 1.09, 95% CI 0.99-1.19 p=0.08; severe wasting, OR: 1.15; 95% CI: 0.32-4.07 p=0.83).

#### Pooled analysis for child underweight

The meta-analysis included data from 45 studies (49 datapoints) with data for moderate underweight (**Figure 4**) and four with data for severe underweight (**Figure 5**). The pooled results for all included studies showed that, compared to adult pregnancy, adolescent pregnancy (≤24 years) was associated with 1.12 times greater odds of moderate underweight (95% CI: 1.00 – 1.26 p=0.04) and 1.21 times greater odds of severe underweight (95% CI: 1.08-1.35 p<0.01) in children 1-59 months.

**Figure 4.**
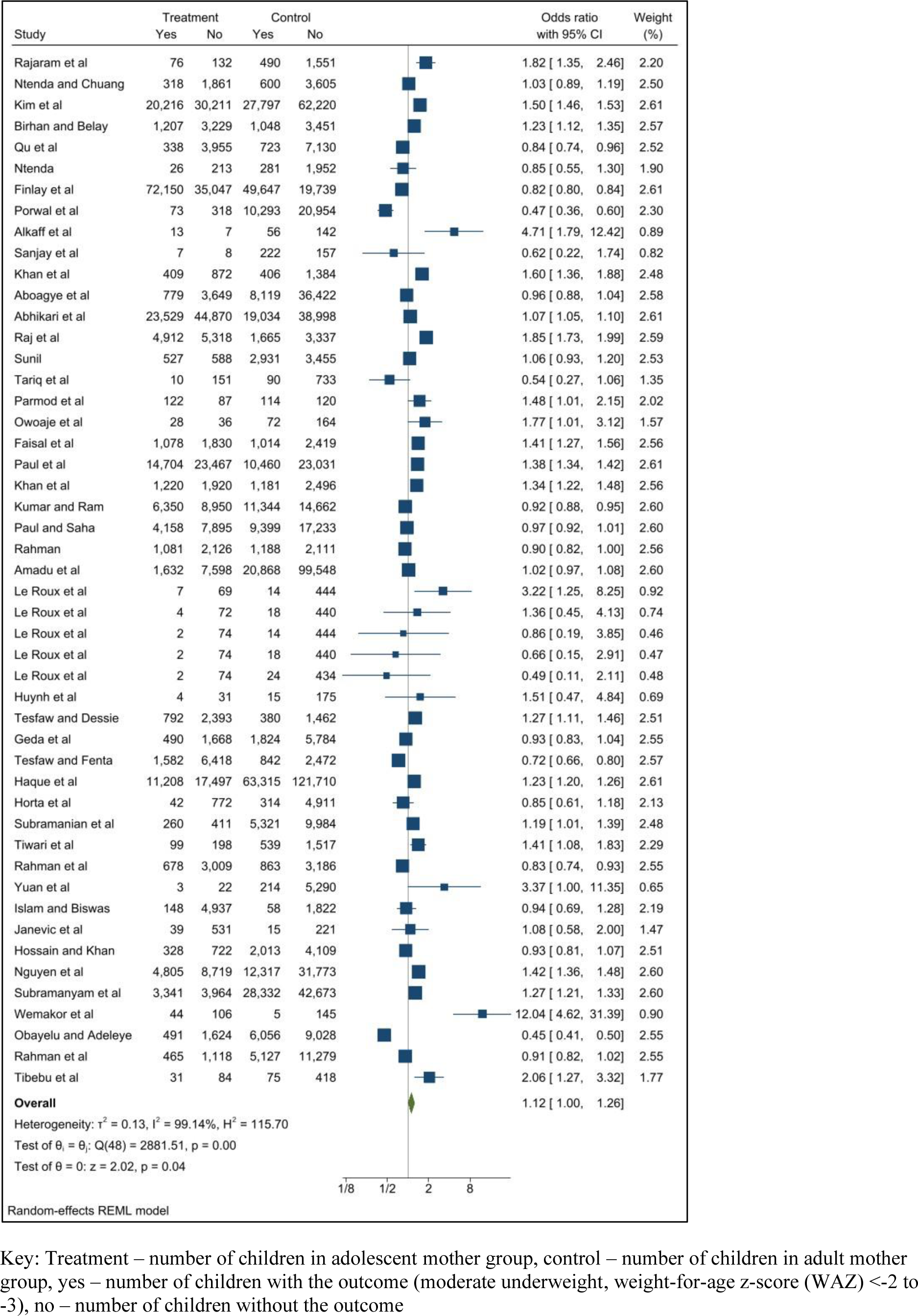
Association between adolescent pregnancy (10-24 years) and the pooled odds of moderate childhood underweight (1-59 months), versus adult pregnancy.

**Figure 5.**
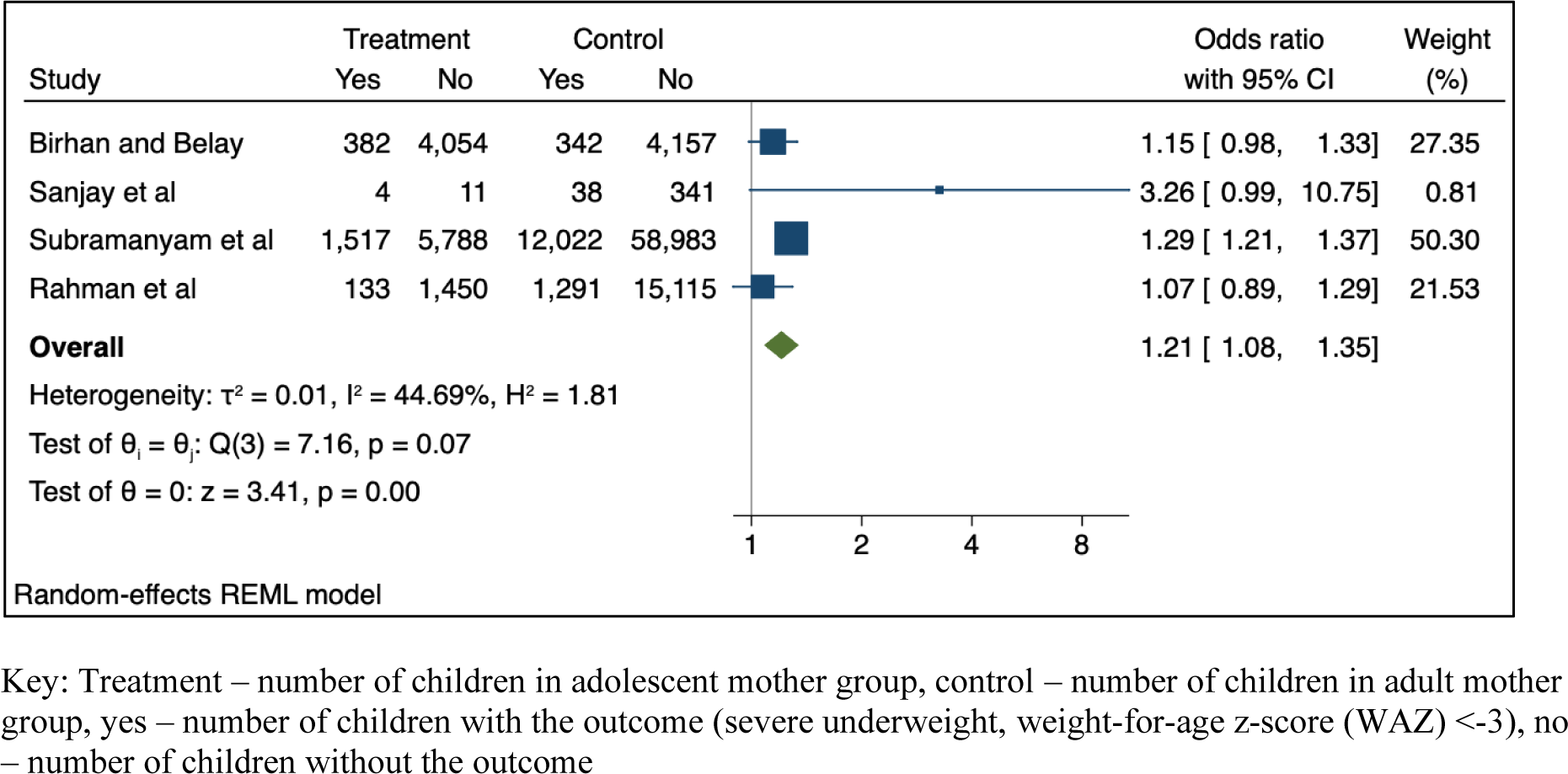
Association between adolescent pregnancy (10-19 years) and the pooled odds of severe childhood underweight (1-59 months), versus adult pregnancy.

Following removal of nine studies for the sensitivity analysis for moderate underweight (**Supplementary Figure 3**), the pooled results for the association with adolescent pregnancy (10-19 years) were no longer statistically significant (OR: 1.09; 95% CI 0.87-1.35 p=0.46). No sensitivity analysis was conducted for severe underweight, as the youngest maternal age category included only adolescents <20 years in all studies.

#### Pooled analysis by geographic location

Given the limited geographic spread of the identified studies, random effect models were only generated for two regions: Asia and SSA.

For Asia, the meta-analysis included 24 studies with data for moderate wasting (**Supplementary Figure 4**) and 28 studies with data for moderate underweight (**Supplementary Figure 5**). The pooled results showed that adolescent pregnancy (≤24 years) was associated with 1.09 times greater odds of moderate wasting (95% CI 1.00–1.19 p=0.04) and 1.16 times greater odds of moderate underweight (95% CI 1.02-1.32 p=0.02) in children 1-59 months compared to adult pregnancy.

Only four studies were included in the random effects model for severe wasting in Asia (**Supplementary Figure 6**), demonstrating no effect of adolescent (≤24 years) compared to adult pregnancy (OR: 1.15; 95% CI 0.32-4.07 p=0.83). Results from the pooled analysis of three studies from Asia (**Supplementary Figure 7**) showed 1.23 times greater odds of severe underweight for children born to adolescent versus adult mothers (95% CI 1.04-1.45 p=0.03).

The random effects models for moderate wasting (**Supplementary Figure 8**) and underweight (**Supplementary Figure 9**) in SSA included 18 and 12 studies (16 data points) respectively. There were no effects of adolescent pregnancy (≤24 years) demonstrated for either outcome (moderate wasting, OR: 1.08; 95% CI 0.93-1.25 p=0.30; and severe wasting, OR: 1.21; 95% CI 0.87-1.67 p=0.26).

Similarly, the random effects models for severe wasting in SSA included only two studies (**Supplementary Figure 10**), with the pooled results showing no effect of adolescent (≤24 years) versus adult pregnancy (OR: 1.03; 95% CI 0.68-2.42 p=0.89). No studies were from SSA presented data for severe underweight.

### Qualitative synthesis

#### Child wasting

Half of the studies (13/26, 50%) included in the qualitative synthesis (**Supplementary Appendix 5a**) reported an association between adolescent versus adult pregnancy and increased risk of moderate wasting in children 1-59 months, compared to 15% (4/26) of studies reporting a reduced risk and 23% (6/26) reporting no difference. Of the two studies with results for severe wasting, one showed no difference in risk for children born to adolescent compared to adult mothers (Ahmed, Ahmed, Roy, Alam, & Hossain, 2012). The other found that, of children receiving treatment for severe wasting, a higher percentage had been born to adolescent mothers (16-20 years) compared to mothers in older age groups (21– 25 years: 32.2%, 26-30 years: 23.1%) (Mokwena & Kachabe, 2022).

#### Child underweight

Most studies (15/24, 63%) which reported data for moderate underweight (**Supplementary Appendix 5b**) indicated an increased risk for children born to adolescent compared to adult mothers (Ahmed et al., 2012; Boah, Azupogo, Amporfro, & Abada, 2019; Friebert, Callaghan-Gillespie, Papathakis, & Manary, 2017; Fuada, Latifah, Yunitawat, & Ashar, 2020; Gbadamosi, Goon, & Tugli, 2017; Hien & Hoa, 2009; Hiruy et al., 2021; Hossain et al., 2020; Ickes, Hurst, & Flax, 2015; Linnemayr, Alderman, & Ka, 2008; Mashal et al., 2008; Nguyen et al., 2017; Poda, Hsu, & Chao, 2017; Schott, Aurino, Penny, & Behrman, 2017; Sobkoviak, Yount, & Halim, 2012). A total of three studies (13%) indicated a reduced risk of underweight for children born to adolescent mothers (Ali et al., 2019; Nakamori et al., 2010) and four (17%) indicated no difference (Akseer et al., 2018; Bekele, Rawstorne, & Rahman, 2021; Hien & Kam, 2008; Kasaye, Bobo, Yilma, & Woldie, 2019; Kumar & Paswan, 2021). Only one study included in the qualitative synthesis presented data for severe underweight and showed no increased risk in children born to adolescent mothers, compared to older mothers (adjusted OR <20 years versus 20-30 years: 1.11 95% CI 0.95-1.29).

#### Outcomes by geographic location

Nine studies in the qualitative synthesis presented data on moderate wasting in Asia (Ahmed et al., 2012; Akseer et al., 2018; Ali et al., 2019; Fuada et al., 2020; Hien & Hoa, 2009; Hien & Kam, 2008; Mashal et al., 2008; Nguyen et al., 2017; Nguyen et al., 2021) and 11 in SSA (Beiersmann et al., 2013; Boah et al., 2019; Friebert et al., 2017; Gbadamosi et al., 2017; Gewa & Yandell, 2012; Ickes et al., 2015; Issah, Yeboah, Kpordoxah, Boah, & Mahama, 2022; Mokwena & Kachabe, 2022; Olodu, Adeyemi, Olowookere, & Esimai, 2019; Poda et al., 2017; Sobkoviak et al., 2012). For Asia, two thirds of studies (6/9) reported an association between adolescent versus adult pregnancy and an increased risk of child wasting compared to approximately a quarter (3/11) of studies for SSA. No comparison by setting was possible for severe wasting, due to the limited number of studies included in the qualitative synthesis for this outcome.

Approximately half of studies from Asia (6/11) (Ahmed et al., 2012; Fuada et al., 2020; Hien & Hoa, 2009; Hossain et al., 2020; Mashal et al., 2008; Nguyen et al., 2017) indicated an increased risk of moderate underweight for children born to adolescent versus adult mothers, compared to two thirds of studies from SSA (8/12) (Boah et al., 2019; Friebert et al., 2017; Gbadamosi et al., 2017; Hiruy et al., 2021; Ickes et al., 2015; Linnemayr et al., 2008; Poda et al., 2017; Sobkoviak et al., 2012). Like severe wasting, a lack of data prevented geographical comparison for severe underweight.

#### Influence of other factors

Of the seven studies that measured maternal nutritional status, 5/7 with maternal BMI measurements observed that a greater proportion of adolescent mothers were underweight than their adult counterparts, or had, on average, lower BMIs (Akseer et al., 2018; R. Das et al., 2022; Nguyen et al., 2017; Nguyen et al., 2021; Nguyen, Scott, Neupane, Tran, & Menon, 2019). In a cohort of adolescent mothers, conditional height-for-age z-score (HAZ) between 12 and 15 years of age was identified as a predictor of WAZ in the offspring between zero and 78 months (β 0.34 p <0.05 in analysis including maternal anthropometry at two time points; β 0.41 p<0.01 for analysis including maternal anthropometry at one time point). This study also found a positive association between dairy consumption in the last 24 hours at age 15 and WAZ in the offspring (β 0.38 p <0.10 in analysis including maternal anthropometry at two time points; β 0.42 p <0.05 for analysis including maternal anthropometry at one time point) (Schott et al., 2017).

In a sub-group analysis of the Mamachiponde RCT in Malawi, in which moderately malnourished women received one of three supplementary foods during pregnancy (Friebert et al., 2017), comparatively more infants born to younger adolescent mothers were underweight at 6- and 12-weeks of age than those born to older adolescent and adult mothers, despite the younger mothers having received a comparatively greater quantity of rations across the study duration.

Of the three studies that assessed the risk of wasting for children of mothers affected by overweight or obesity, two showed no effect of maternal age (S. Das et al., 2019; Masibo, Humwa, & Macharia, 2020) and one indicated a reduced risk of wasting for children born to adolescent mothers, compared to their older counterparts (adjusted OR: wasting: <20 years: reference group, 20 – 29 years: 2.56 (2.11 - 3.1) p <0.005, 30 – 39 years: 3.67 (3.03 - 4.46) p <0.005, 40 – 49 years: 3.90 (3.14 - 4.85) p <0.005) (Biswas, Townsend, Magalhaes, Hasan, & Mamun, 2021). Findings for the risk of underweight were similar: one study showed no effect of maternal age (Masibo et al., 2020); one indicated a reduced risk of underweight for children born to mothers 21-25 years (adjusted OR: 2.16 (1.08 - 4.48) p <0.05) (S. Das et al., 2019); and one indicated a reduced risk of underweight in children born to adolescent mothers compared to those born to mothers in all adult age groups (adjusted OR: <20 years: reference group, 20 – 29 years: 2.56 (2.11 - 3.09) p <0.005, 30 – 39 years: 3.65 (3.01 - 4.43) p <0.005, 40 – 49 years: 3.87 (3.12 - 4.80) p <0.005) (Biswas et al., 2021).

Studies which explored the influence of social factors, suggested that, for adolescent mothers, poverty and low educational attainment may be associated with a lack of knowledge about childcare and feeding practices and lower hygiene standards which, in turn, increases the risk of diarrhoea and malnutrition during childhood (Fuada et al., 2020). Child marriage was associated with reduced autonomy and access to healthcare for both the mother and child (Mashal et al., 2008; Paul, Chouhan, & Zaveri, 2019; Raj et al., 2010).

## Discussion

This review explored the associations between adolescent pregnancy and the risk of childhood wasting and underweight. Results from the meta-analysis showed that children born to mothers ≤24 years were at greater risk of moderate and severe underweight than those born to adult mothers. Pooled estimates showed no statistically significant associations between adolescent pregnancy and risk of childhood wasting. However, results from individual studies were mixed and stratified analyses indicated a higher risk of moderate wasting in children born to adolescent versus adult mothers in Asia. These findings were largely supported by the qualitative synthesis, where most studies found an association between adolescent pregnancy and risk of childhood underweight (moderate and severe). Evidence of an association with risk of wasting was less consistent.

Previous evidence has demonstrated a protective effect of older (20-35 years) compared to younger (<20 years) maternal age on risk of childhood underweight, but not wasting (Kerac, Frison, Connell, Page, & McGrath, 2019; Mertens, Benjamin-Chung, Colford, Coyle, et al., 2020). Associations between adolescent pregnancy and risk of childhood stunting have also been established, with low birthweight shown to mediate this relationship (Maravilla, Betts, Adair, & Alati, 2020). For mothers who experience repeated early pregnancies and increased nutritional deficits, these associations may be further exacerbated (Maravilla et al., 2020). Underweight is a composite indicator of wasting and stunting (Sadler, Khara, & Sessions, 2021) which may better represent persistent and repeated episodes of growth faltering across the pregnancy-postpartum continuum and thus, the longer-term risks associated with early pregnancy/motherhood across childhood. Since children who are underweight (and, by nature, simultaneously wasted and stunted) are at comparatively greater risk of mortality than those with individual deficits (wasting or stunting alone) (Myatt et al., 2018), further exploration into the relationship between adolescent pregnancy and risk of underweight as children age is needed. This may, in turn, provide a key target for interventions aiming to mitigate the persistent burden of child undernutrition in LMICs, particularly in contexts where the rate of adolescent pregnancy is high.

In addition to the finding for moderate wasting, results from the stratified analysis showed that children born to adolescent (≤24 years) mothers were at a greater risk of moderate and severe underweight than those born to adult mothers in Asia. There were no significant pooled effects of adolescent pregnancy on any child outcomes when analyses were run using data from SSA. While this may, in part, reflect the comparatively greater availability of data for Asia, it also suggests contextual variation in the biological, social and/or cultural factors which influence the relationship between maternal age and offspring growth and development. For example, in contrast to the pooled findings for Asia, one study from Vietnam, found no association between adolescent pregnancy and risk of underweight in the offspring – attributing these findings to high levels of familial involvement in the care of infants born to adolescent mothers in this context which likely reduces social and/or socioeconomic impacts of adolescent motherhood on child growth outcomes (Nakamori et al., 2010). This highlights the need for contextualised approaches that aim to minimise inequalities to both prevent early marriage and pregnancy, and support optimal nutrition, health and development of adolescent girls, and their offspring, should they become pregnant (Lelijveld et al., 2023).

Previous research supports the contribution of social determinants of health, including ethnicity, socioeconomic status and education, to the risk of adverse pregnancy outcomes in adolescent mothers (Amjad et al., 2019). While this review aimed to explore the influence of other biological and/or social factors in the association between adolescent pregnancy and wasting and/or underweight as children age, a dearth of evidence restricted our ability to do so. Most studies only presented unadjusted rather than adjusted associations between adolescent age and subsequent child undernutrition – this precluded statistical examination of the role of other factors which likely act as confounders or effect modifiers. In addition, in cases where pooled estimates were significant, effect sizes were small and may have disappeared if such factors were accounted for – highlighting that, while adolescent pregnancy may be a useful proxy of risk, there is a need to focus on a wider group of at-risk mothers. For example, limited data on maternal nutritional status suggested that adolescent mothers were more likely than their adult counterparts to have lower BMIs or to be underweight and that maternal short stature may influence the risk of underweight in children (0-6 years) born to adolescent mothers. This could, at least in part, explain the association between adolescent pregnancy and childhood underweight; it may also explain why studies showed mixed results since adolescent pregnancy may be a proxy for maternal nutritional status (and potentially other maternal factors), rather than a risk factor itself.

Some studies demonstrated effects of child marriage on lack of access to healthcare services (Mashal et al., 2008; Paul et al., 2019; Raj et al., 2010) and reflected on the potential role of poverty and low educational attainment on adolescents’ infant feeding and care practices (Fuada et al., 2020). However, exploration into the potential impacts on child anthropometric outcomes was lacking and more research is needed to understand the pathways between adolescent pregnancy/motherhood and child underweight and wasting across contexts. This will help to inform development of tailored interventions and services which support optimal growth and development of both adolescent girls and their offspring through pregnancy and postpartum periods. While interventions such as pre- and postnatal supplementation show promise – with documented benefits on birth size and on linear growth up to 12 months of age – evidence of benefits on the risk of wasting and underweight in later childhood is lacking, as is evidence comparing potential benefits for adolescent and adult mothers (Argaw et al., 2023).

### Strengths and limitations

The systematic approach, broad geographical coverage and large number of papers included in this review are notable strengths. A more inclusive definition of adolescence was also taken, with a sensitivity analysis used to explore the data in depth. However, several limitations are also acknowledged.

First, many of the included studies were cross-sectional and utilised open-source datasets. While efforts were made to catalogue and cross-check the datasets, and remove direct duplications, there is a risk that one or more datasets were over-represented in the meta-analysis and this may have affected the reliability of the pooled effect estimates (Senn, 2009). Similarly, some countries (e.g., Bangladesh and India) were over-represented in the data, and may have influenced the comparatively strong associations observed in Asia. Since social and cultural factors are likely to influence the relationships between adolescent pregnancy and offspring growth and development, over- or under-representation from countries/regions may lead to an over- or under-reporting of the true effect. To consider these potential differences, sub-analyses by region were intended, but could only be conducted for SSA and Asia due to a lack of data from other regions, limiting the conclusions that can be drawn. It is possible that restricting inclusion of studies to those published in English may have contributed to the under-representation of studies from certain geographical regions such as North Africa and Latin America.

This review focused only on wasting and underweight since these are the two forms of severe malnutrition most amenable to short-term change; stunting has closely related but separate aetiology and is more difficult to address (Kerac et al., 2020). There is an extensive body of literature focused on stunting and this would warrant its own stand-alone review. It is however possible that an association between adolescent pregnancy and stunting mediates some of our observations: if for example adolescent pregnancy affects length as well as weight this might partly explain our observation of greater apparent association with weight-for-age (underweight) but not so strong an association with weight-for-length (wasting). There was substantial heterogeneity in the study designs, particularly related to the categorisation of maternal age, which challenged the comparability between studies and may have diluted the effect sizes in the meta-analysis. However, consideration of this was addressed, at least in part, via inclusion of a sensitivity analysis which compared the results of the fixed effects models when adolescence was defined as ≤24 years versus 10-19 years. As previously mentioned, outcome data was rarely disaggregated according to child age, limiting our ability to assess differences in risk across life stages (<6 months, 6-<24 months, 24-59 months).

The random effects models created for the meta-analysis utilised prevalence data to estimate pooled ORs, predominantly from observational studies. Thus, there is an intrinsic risk of confounding in the statistical analysis, which must be considered when interpreting the results. Evidence suggests that other factors, e.g., care practices, and socioeconomic factors, mediate the relationships between adolescent pregnancy and both low birth weight and childhood stunting and, therefore may be influential in the associations with risk of wasting and underweight (Fall et al., 2015; Yu, Mason, Crum, Cappa, & Hotchkiss, 2016).

## Conclusion

This systematic review and meta-analysis provided evidence that being born to an adolescent, compared to an adult, mother is associated with increased risk of childhood underweight. Pooled analyses showed no overall associations between adolescent pregnancy and risk of childhood wasting; however, there was heterogeneity in individual studies and stratified analyses indicated an increased risk of moderate wasting for children born to mothers ≤24 years in Asia. While evidence on the potential role of biological/social factors was limited, available studies suggest an important potential intermediary role of maternal nutritional status. This could be important for future interventions and warrants further exploration. Particularly in contexts where rates of adolescent pregnancy remain high, interventions to both delay adolescent pregnancy and improve adolescent nutritional status could help reduce the risk of undernutrition in children and contribute to breaking the intergenerational cycle of malnutrition.

## Data Availability

All data produced in the present study are available upon reasonable request to the authors

## Acknowledgements

The authors were able to undertake this work due to the generous support of UNICEF. MK gratefully acknowledges funding from the Eleanor Crook Foundation for related work on infant/maternal malnutrition. The ideas, opinions and comments included are entirely the responsibility of the article’s authors and do not necessarily represent or reflect the policies of the donor.

## Author contributions

CW, SVW, NL and MK designed the research. CW conducted the literature search and meta-analysis. CW and CKW conducted the article screening, with review support by SVW. CW and SVW wrote the paper. All authors contributed to interpretation of results and reviewed the final version for submission.

