## Supplementary Information for "Adolescent pregnancy is associated with child undernutrition: systematic review and meta-analysis"

### Supplementary Appendix 1. Search strategy and results

1 (Adolescen* or teen* or youth* or young person* or young people* or "10-19 year*").mp. [mp=title, book title, abstract, original title, name of substance word, subject heading word, floating sub-heading word, keyword heading word, organism supplementary concept word, protocol supplementary concept word, rare disease supplementary concept word, unique identifier, synonyms] (2306108)
2 adolescent/ or young adult/ or child/ (3518101)
3 1 or 2 (3619539)
4 (Pregnan* or mother* or matern*).mp. [mp=title, book title, abstract, original title, name of substance word, subject heading word, floating sub-heading word, keyword heading word, organism supplementary concept word, protocol supplementary concept word, rare disease supplementary concept word, unique identifier, synonyms] (1330001)
5 Pregnant Women/ (12699)
6 pregnancy/ or pregnancy in adolescence/ (957946)
7 maternal age/ (20750)
8 Adolescent Mothers/ (46)
9 4 or 5 or 6 or 7 or 8 (1330001)
10 Nutrition* status.mp. [mp=title, book title, abstract, original title, name of substance word, subject heading word, floating sub-heading word, keyword heading word, organism supplementary concept word, protocol supplementary concept word, rare disease supplementary concept word, unique identifier, synonyms] (74538)
11 malnutrition.mp. or *nutrition/ or *weight/ or malnourished.mp. (61251)
12 Nutritional Status/ (51802)
13 malnutrition/ or deficiency diseases/ (25335)
14 severe acute malnutrition/ or kwashiorkor/ (3030)
15 Starvation/ (10139)
16 overnutrition/ or overweight/ or obesity/ or obesity, maternal/ (220192)
17 Wasting Syndrome/ (1432)
18 protein deficiency/ or protein-energy malnutrition/ (12271)
19 Thinness/ (7299)
20 10 or 11 or 12 or 13 or 14 or 15 or 16 or 17 or 18 or 19 (356466)
21 9 or 20 (1653395)
22 (Infant* or child* or baby or babies or toddler* or offspring or next generation or pre-school child or "Under 5 years").mp. [mp=title, book title, abstract, original title, name of substance word, subject heading word, floating sub-heading word, keyword heading word, organism supplementary concept word, protocol supplementary concept word, rare disease supplementary concept word, unique identifier, synonyms] (3404648)
23 child/ or child, preschool/ or infant/ (2354343)
24 22 or 23 (3404648)
25 (Wasting or wasted or kwashiorkor or marasmus or "weight for height" or WHZ or Growth falter* or underweight or "weight for age" or WAZ or nutrition* depriv* or starvation or famine* or acute malnutrition or moderate malnutrition).mp. [mp=title, book title, abstract, original title, name of substance word, subject heading word, floating sub-heading word, keyword heading word, organism supplementary concept word, protocol supplementary concept word, rare disease supplementary concept word, unique identifier, synonyms] (107973)
26 exp malnutrition/ (132030)
27 (afghanistan or albania or algeria or american samoa or angola or "antigua and barbuda" or antigua or barbuda or argentina or armenia or armenian or aruba or azerbaijan or bahrain or bangladesh or barbados or republic of belarus or belarus or byelarus or belorussia or byelorussian or belize or british honduras or benin or dahomey or bhutan or bolivia or "bosnia and herzegovina" or bosnia or herzegovina or botswana or bechuanaland or brazil or brasil or bulgaria or burkina faso or

burkina fasso or upper volta or burundi or urundi or cabo verde or cape verde or cambodia or kampuchea or khmer republic or cameroon or cameron or cameroun or central african republic or ubangi shari or chad or chile or china or colombia or comoros or comoro islands or iles comores or mayotte or democratic republic of the congo or democratic republic congo or congo or zaire or costa rica or "cote d'ivoire" or "cote d' ivoire" or cote divoire or cote d ivoire or ivory coast or croatia or cuba or cyprus or czech republic or czechoslovakia or djibouti or french somaliland or dominica or dominican republic or ecuador or egypt or united arab republic or el salvador or equatorial guinea or spanish guinea or eritrea or estonia or eswatini or swaziland or ethiopia or fiji or gabon or gabonese republic or gambia or "georgia (republic)" or georgian or ghana or gold coast or gibraltar or greece or grenada or guam or guatemala or guinea or guinea bissau or guyana or british guiana or haiti or hispaniola or honduras or hungary or india or indonesia or timor or iran or iraq or isle of man or jamaica or jordan or kazakhstan or kazakh or kenya or "democratic people's republic of korea" or republic of korea or north korea or south korea or korea or kosovo or kyrgyzstan or kirghizia or kirgizstan or kyrgyz republic or kirghiz or laos or lao pdr or "lao people's democratic republic" or latvia or lebanon or lebanese republic or lesotho or basutoland or liberia or libya or libyan arab jamahiriya or lithuania or macau or macao or republic of north macedonia or macedonia or madagascar or malagasy republic or malawi or nyasaland or malaysia or malay federation or malaya federation or maldives or indian ocean islands or indian ocean or mali or malta or micronesia or federated states of micronesia or kiribati or marshall islands or nauru or northern mariana islands or palau or tuvalu or mauritania or mauritius or mexico or moldova or moldovian or mongolia or montenegro or morocco or ifni or mozambique or portuguese east africa or myanmar or burma or namibia or nepal or netherlands antilles or nicaragua or niger or nigeria or oman or muscat or pakistan or panama or papua new guinea or new guinea or paraguay or peru or philippines or philipines or phillipines or phillippines or poland or "polish people's republic" or portugal or portuguese republic or puerto rico or romania or russia or russian federation or ussr or soviet union or union of soviet socialist republics or rwanda or ruanda or samoa or pacific islands or polynesia or samoan islands or navigator island or navigator islands or "sao tome and principe" or saudi arabia or senegal or serbia or seychelles or sierra leone or slovakia or slovak republic or slovenia or melanesia or solomon island or solomon islands or norfolk island or norfolk islands or somalia or south africa or south sudan or sri lanka or ceylon or "saint kitts and nevis" or "st. kitts and nevis" or saint lucia or "st. lucia" or "saint vincent and the grenadines" or saint vincent or "st. vincent" or grenadines or sudan or suriname or surinam or dutch guiana or netherlands guiana or syria or syrian arab republic or tajikistan or tadjikistan or tadzhikistan or tadzhik or tanzania or tanganyika or thailand or siam or timor leste or east timor or togo or togolese republic or tonga or "trinidad and tobago" or trinidad or tobago or tunisia or turkey or turkmenistan or turkmen or uganda or ukraine or uruguay or uzbekistan or uzbek or vanuatu or new hebrides or venezuela or vietnam or viet nam or middle east or west bank or gaza or palestine or yemen or yugoslavia or zambia or zimbabwe or northern rhodesia or global south or africa south of the sahara or sub-saharan africa or subsaharan africa or africa, central or central africa or africa, northern or north africa or northern africa or magreb or maghrib or sahara or africa, southern or southern africa or africa, eastern or east africa or eastern africa or africa, western or west africa or western africa or west indies or indian ocean islands or caribbean or central america or latin america or "south and central america" or south america or asia, central or central asia or asia, northern or north asia or northern asia or asia, southeastern or southeastern asia or south eastern asia or southeast asia or south east asia or asia, western or western asia or europe, eastern or east europe or eastern europe or developing country or developing countries or developing nation? or developing population? or developing world or less developed countr* or less developed nation? or less developed population? or less developed world or lesser developed countr* or lesser developed nation? or lesser developed population? or lesser developed world or under developed countr* or under developed nation? or under developed population? or under developed world or underdeveloped countr* or underdeveloped nation? or underdeveloped population? or underdeveloped world or middle income countr* or middle income nation? or middle income population? or low income countr* or low income nation? or low income population? or lower income countr* or lower income nation? or lower income population? or underserved countr* or underserved nation? or underserved population? or underserved world or under served countr* or under served nation? or under served population? or under served world or deprived countr* or deprived nation? or deprived population? or deprived world or poor countr* or poor nation? or poor population? or poor world or poorer countr* or poorer nation? or poorer population? or poorer world or developing econom* or less developed econom* or lesser developed econom* or under developed econom* or underdeveloped econom* or middle income econom* or low income econom* or lower income econom* or low gdp or low gnp or low gross domestic or low gross national or lower gdp or lower gnp or lower gross domestic or lower gross national or lmic or lmics or third world or lami countr* or transitional countr* or emerging economies or emerging nation?).mp. [mp=title, book title, abstract, original title, name of substance word, subject heading word, floating sub-heading word, keyword heading word, organism supplementary concept word, protocol supplementary concept word, rare disease supplementary concept word, unique identifier, synonyms] (2317513)

28 25 or 26 (222101)

29 limit 28 to yr="1990 -Current" (156748)

30 3 and 21 and 24 and 27 and 28 and 29 (6764)

*Note: numbers in parenthesis show the number of results per line*

### Supplementary Appendix 2. Study characteristics

| Ref. | Publication Reference | Country | Data source | Sample size | Child age category* (months) | Wasting outcome | | Underweight outcome | | Youngest maternal age <20 years | Data extractable for meta-analysis | Quality assessment |
|---|---|---|---|---|---|---|---|---|---|---|---|---|
| | | | | | | Mod[a] | Sev[b] | Mod[c] | Sev[d] | | | |
| **Cross-sectional studies** | | | | | | | | | | | | |
| 29 | (Adhikari et al., 2021) | India | NFHS-4 2015-16 | 126431 | 0-36 | ■ | ⊕ | ■ | ⊕ | ⊕ | ■ | Good |
| 28 | (Aboagye et al., 2021) | 32 Sub-Saharan Africa countries | DHS data 2010-2020 | 48968 | 6-23 | ■ | ⊕ | ■ | ⊕ | ■ | ■ | Good |
| 27 | (Ahmed, Ahmed, Roy, Alam, & Hossain, 2012) | Bangladesh | Bangladesh National Nutrition Programme baseline survey 2004 | 8858 | 0-24 | ■ | ■ | ■ | ■ | ■ | ⊕ | Fair |
| 46 | (Akseer et al., 2018) | Afghanistan | ANNS 2013 | 14000 | 0-59 | ■ | ⊕ | ■ | | ■ | ⊕ | Good |
| 14 | (Ali et al., 2019) | Bangladesh | Nobokoli programme baseline data | 6468 | 0-59 | ■ | ⊕ | ■ | ⊕ | ⊕ | ⊕ | Fair |
| 19 | (Alkaff, Flynn, Sukmajaya, & Salamah, 2020) | Indonesia | Cross-sectional survey and secondary data from child health records | 218 | 0-59 | ⊕ | ⊕ | ■ | ⊕ | ■ | ■ | Good |
| 53 | (Amadu et al., 2021) | 31 Sub-Saharan countries | DHS 2010-2019 | 129646 | 0-59 | ■ | ⊕ | ■ | ⊕ | ■ | ■ | Good |
| 22 | (Amare, Ahmed, & Mehari, 2019) | Ethiopia | DHS 2016 | 9419 | 0-59 | ■ | ⊕ | ⊕ | ⊕ | ■ | ■ | Good |
| 56 | (Beiersmann et al., 2013) | Burkina Faso | Cross-sectional data | 460 | 6-51 | ■ | ⊕ | ■ | ⊕ | | ⊕ | Good |
| 86 | (Bekele, Rawstorne, & Rahman, 2021) | Ethiopia | DHS 2000 and 2016 | 21514 | 0-59 | ■ | ⊕ | ■ | ⊕ | ⊕ | ⊕ | Good |
| 9 | (Birhan & Belay, 2021) | Ethiopia | DHS 2016 | 8935 | 0-59 | ⊕ | ⊕ | ■ | ■ | ■ | ■ | Fair |
| 70 | (Biswas, Townsend, Magalhaes, Hasan, & Mamun, 2021) | South and South-East Asian countries | DHS 2007-2016 | 798961 | 0-59 | ■ | ⊕ | ■ | ⊕ | ■ | ⊕ | Good |
| 36 | (Boah, Azupogo, Amporfro, & Abada, 2019) | Ghana | DHS 2014 | 2636 | 0-59 | ■ | ⊕ | ■ | ⊕ | ■ | ⊕ | Good |
| 78 | (S. Das et al., 2019) | Bangladesh | DHS 2014 | 5951 | 0-59 | ■ | ⊕ | ■ | ⊕ | ■ | ⊕ | Good |
| 20 | (Fagbamigbe, Kandala, & Uthman, 2020) | 51 LMIC | DHS data 2010-2018 | 532680 | 0-59 | ⊕ | ■ | ⊕ | ⊕ | | ■ | Good |
| 47 | (Faisal Ahmed et al., 2021) | Bangladesh | DHS 2014 | 6341 | 0-59 | ■ | ⊕ | ■ | ⊕ | ■ | ■ | Good |
| 16 | (Finlay, Ozaltin, & Canning, 2011) | 55 LMIC | 118 DHS surveys 1990-2008 | 176583 | 0-59 | ■ | ⊕ | ■ | ⊕ | ■ | ■ | Good |

| | | | | | | | | | | | | |
|---|---|---|---|---|---|---|---|---|---|---|---|---|
| 8 | (Fuada, Latifah, Yunitawat, & Ashar, 2020) | Indonesia | Basic health research data 2013 | 978 | 0-59 | ■ | ⊕ | ■ | ⊕ | ■ | ⊕ | Fair |
| 81 | (Gbadamosi, Goon, & Tugli, 2017) | South Africa | Cross-sectional data from the MAL-ED study | 186 | 1-12 | ■ | ⊕ | ■ | ⊕ | ■ | ⊕ | Good |
| 60 | (Geda et al., 2021) | Ethiopia | DHS 2016 | 9218 | 6-59 | ■ | ⊕ | ■ | ⊕ | ⊕ | ■ | Good |
| 88 | (Gewa & Yandell, 2012) | Kenya | DHS survey 2003 | 1851 | 0-60 | ■ | ⊕ | ■ | ⊕ | ⊕ | ⊕ | Good |
| 84 | (Ghimire, Aryal, Gupta, & Sapkota, 2020) | Nepal | Cross-sectional data from children admitted to the OTP | 398 | 6-59 | ⊕ | ■ | ⊕ | ⊕ | ■ | ■ | Good |
| 62 | (Haque et al., 2022) | South and South-East Asian Countries | DHS 2014-2018 | 213730 | 0-59 | ■ | ⊕ | ■ | ⊕ | ■ | ■ | Good |
| 64 | (Hien & Hoa, 2009) | Vietnam | Cross-sectional data | 383 | 6-36 | ■ | ⊕ | ■ | ⊕ | ⊕ | ⊕ | Good |
| 65 | (Hien & Kam, 2008) | Vietnam | Cross-sectional data | 650 | 0-59 | ■ | ⊕ | ■ | ⊕ | ⊕ | ⊕ | Good |
| 13 | (Hiruy et al., 2021) | Ethiopia | DHS 2000-2016 | 8003 | 6-23 | ■ | ⊕ | ■ | ⊕ | ■ | ⊕ | Good |
| 66 | (Horta et al., 2013) | Brazil | National Survey of Indigenous People's Health and Nutrition 2008-2009 | 6075 | 0-59 | ■ | ⊕ | ■ | ⊕ | ■ | ■ | Good |
| 31 | (F. B. Hossain et al., 2020) | Bangladesh, India, Pakistan, Maldives and Nepal | DHS 2009 - 2016 | 146996 | 24-59 | ⊕ | ⊕ | ■ | ⊕ | ■ | ⊕ | Good |
| 83 | (M. B. Hossain & Khan, 2018) | Bangladesh | DHS 2014 | 7173 | 0-59 | ■ | ⊕ | ■ | ⊕ | ■ | ■ | Good |
| 55 | (Huynh, Huynh, Nguyen, Do, & Khanh Tran, 2019) | Vietnam | Cross-sectional data | 225 | 6-59 | ⊕ | ⊕ | ■ | ⊕ | ⊕ | ■ | Good |
| 57 | (Ickes, Hurst, & Flax, 2015) | Uganda | DHS 2006 and 2011 | 1009, 888 | 0-23 | ■ | ⊕ | ■ | ⊕ | ■ | ⊕ | Good |
| 76 | (Islam & Biswas, 2020) | Bangladesh | DHS 2014 | 6965 | 0-59 | ■ | ⊕ | ■ | ⊕ | ■ | ■ | Good |
| 10 | (Issah, Yeboah, Kpordoxah, Boah, & Mahama, 2022) | Nigeria | DHS 2018 | 2425 | 0-59 | ■ | ⊕ | ⊕ | ⊕ | ⊕ | ⊕ | Good |
| 82 | (Janevic, Petrovic, Bjelic, & Kubera, 2010) | Serbia | MICS 2005 | 1192 | 0-59 | ■ | ⊕ | ■ | ⊕ | ■ | ■ | Good |
| 1 | (Karim et al., 2021) | Bangladesh | Study data | 540 | 6-59 | ■ | ⊕ | | ⊕ | ■ | ■ | Good |
| 72 | (Kasaye, Bobo, Yilma, & Woldie, 2019) | Ethiopia | DHS 2016 | 9494 | 0-59 | ■ | ⊕ | ■ | ⊕ | ⊕ | ■ | Good |
| 26 | (S. Khan, Zaheer, & Safdar, 2019) | Pakistan | DHS 2012-2013 | 3071 | 0-59 | ■ | ⊕ | ■ | ⊕ | ■ | ■ | Good |
| 49 | (M. S. Khan, Halder, Rashid, Afroja, & Islam, 2020) | Bangladesh | DHS 2014 | 8092 | 0-59 | ⊕ | ⊕ | ■ | ⊕ | ■ | ■ | Good |
| 7 | (Kim et al., 2019) | India | INFHS-4 2015-16 | 140,444 | 0-59 | ■ | ⊕ | ■ | ⊕ | ■ | ■ | Good |

| | | | | | | | | | | | | |
|---|---|---|---|---|---|---|---|---|---|---|---|---|
| 18 | (R. Kumar & Paswan, 2021) | India | NFHS-3 2005-2006, and NFHS-4 2015-2016 | 11858 (NFHS-3), 92630 (NFHS-4) | 0-59 | ⊕ | ⊕ | ■ | ⊕ | ■ | ⊕ | Good |
| 50 | (A. Kumar & Ram, 2013) | India | NFHS 2005-2006 | 41306 | 0-59 | ⊕ | ⊕ | ■ | ⊕ | ⊕ | ■ | Good |
| 77 | (R. Kumar, Abbas, Mahmood, & Somrongthong, 2019) | Pakistan | MICS 2014 | 24042 | 0-59 | ⊕ | ⊕ | ■ | ■ | ■ | ■ | Good |
| 21 | (Linnemayr, Alderman, & Ka, 2008) | Senegal | Baseline data: nutrition intervention programme | 4296 | 0-35 | ⊕ | ⊕ | ■ | ⊕ | ⊕ | ⊕ | Good |
| 40 | (Mashal et al., 2008) | Afghanistan | Cross-sectional survey | 2472 | 0-59 | ■ | ⊕ | ■ | ⊕ | ■ | ⊕ | Good |
| 32 | (Masibo, Humwa, & Macharia, 2020) | Kenya | DHS 2014 | 7830 | 0-59 | ■ | ⊕ | ■ | ⊕ | ■ | ⊕ | Good |
| 37 | (Mena-Melendez, 2020) | Latin America: Bolivia, Colombia, Guatemala and Peru | DHS 1986-2015 | 15827 | 0-59 | ■ | ⊕ | ⊕ | ⊕ | ■ | ⊕ | Good |
| 80 | (Mokwena & Kachabe, 2022) | South Africa | Cross-sectional data | 94 | 1-36 | ■ | ■ | ⊕ | ⊕ | ■ | ⊕ | Fair |
| 44 | (Mutunga, Frison, Rava, & Bahwere, 2020) | Cambodia, Lao PDR, Myanmar, Thailand, Timor-Leste, Vietnam | DHS 2014, Lao PDR MICS 2017, Myanmar DHS 2015, Thailand MICS 2015/16, Timor-Leste NFNS 2013, and Vietnam MICS 2011 | 42308 | 0-59 | ■ | ⊕ | ⊕ | ⊕ | ■ | ■ | Good |
| 68 | (Nakamori et al., 2010) | Vietnam | Cross-sectional data | 188 | 6-18 | ⊕ | ⊕ | ■ | ⊕ | ⊕ | ⊕ | Fair |
| 2 | (Nguyen et al., 2021) | Bangladesh | DHS 1996-2017 | 12006 | 0-59 | ■ | ⊕ | ⊕ | ⊕ | ■ | ⊕ | Good |
| 63 | (Nguyen et al., 2017) | Bangladesh | Baseline data: scaled up MNCH programme | 2000 | 0-5 | ■ | ⊕ | ■ | ⊕ | ■ | ⊕ | Good |
| 85 | (Nguyen, Scott, Neupane, Tran, & Menon, 2019) | India | NFHS-4 2015-16 | 60096 | 0-59 | ■ | ⊕ | ■ | ⊕ | ■ | ■ | Good |
| 6 | (Ntenda & Chuang, 2018) | Malawi | DHS 2004 and 2010 | 6384 | 0-59 | ■ | ⊕ | ■ | ⊕ | ⊕ | ■ | Good |
| 15 | (Ntenda, 2019) | Malawi | DHS 2015-16 | 6033 | 0-59 | ■ | ⊕ | ■ | ⊕ | ⊕ | ■ | Good |
| 90 | (Obayelu & Adeleye, 2021) | Nigeria | DHS 2013 | 17199 | 0-59 | ■ | ⊕ | ⊕ | ⊕ | ■ | ■ | Good |
| 67 | (Olodu, Adeyemi, Olowookere, & Esimai, 2019) | Nigeria | Cross-sectional survey using | 300 | 6-59 | ■ | ⊕ | ■ | ⊕ | ■ | ⊕ | Good |

| | | | | | | | | | | | | |
|---|---|---|---|---|---|---|---|---|---|---|---|---|
| | | | primary health care centres records | | | | | | | | | |
| 42 | (Pramod Singh, Nair, Grubesic, & Connell, 2009) | Nepal | Cross-sectional survey | 443 | 6-36 | ⊕ | ⊕ | ■ | ⊕ | ■ | ■ | Good |
| 48 | (Paul, Chouhan, & Zaveri, 2019) | India | NFHS-4 2015-16 | 80539 | 0-59 | ■ | ⊕ | ■ | ⊕ | ■ | ■ | Good |
| 51 | (Paul & Saha, 2022) | India | NFHS-4 2015-16 | 38685 | 0-59 | ■ | ⊕ | ■ | ⊕ | ⊕ | ■ | Good |
| 39 | (Poda, Hsu, & Chao, 2017) | Burkina Faso | DHS 2010 | 6337 | 0-59 | ■ | ⊕ | ■ | ⊕ | ■ | ⊕ | Good |
| 17 | (Porwal et al., 2021) | India | Comprehensive National Nutrition Survey | 35452 | 0-59 | ■ | ⊕ | ■ | ⊕ | ■ | ■ | Good |
| 12 | (Qu et al., 2017) | China | Cross sectional survey | 12146 | 6-35 | ■ | ⊕ | ■ | ⊕ | | ■ | Good |
| 91 | (A. Rahman & Hossain, 2022) | Bangladesh | DHS 2014 | 17989 | 0-59 | ■ | ■ | ■ | ■ | ■ | ■ | Good |
| 52 | (M. M. Rahman, 2015) | Bangladesh | BDHS 2011 | 6506 | 0-59 | ■ | ⊕ | ■ | ⊕ | ⊕ | ■ | Good |
| 73 | (M. A. Rahman, Halder, Rahman, & Parvez, 2021) | Bangladesh | DHS 2017-18 | 7738 | 0-59 | ■ | ⊕ | ■ | ⊕ | ⊕ | ■ | Good |
| 79 | (M. S. Rahman, Rahman, Maniruzzaman, & Howlader, 2020) | Bangladesh | DHS 2014 | 7131 | 0-59 | ■ | ■ | ⊕ | ⊕ | ■ | ■ | Good |
| 33 | (Raj et al., 2010) | India | NFHS 2005-2006 | 19392 | 0-59 | ■ | ⊕ | ■ | ⊕ | ■ | ■ | Good |
| 5 | (Schott, Aurino, Penny, & Behrman, 2017) | India: Kerala and Goa | NFHS-I 1995 | 2249 | 0-59 | ■ | ⊕ | ■ | ⊕ | ■ | ■ | Good |
| 38 | (Rodgers, Kim, & Subramanian, 2020) | India | NFHS-4 2015-16 | 139116 | 6-59 | ■ | ⊕ | ■ | ⊕ | ■ | ⊕ | Good |
| 45 | (Samuel et al., 2022) | Ethiopia, | Baseline data: effectiveness study | 2036 | 6-11 | ■ | ⊕ | ⊕ | ⊕ | ⊕ | ⊕ | Good |
| 23 | (Sanjay, Gaurav, Suman, & Ramchandra, 2020) | Rajasthan | Cross-sectional survey | 396 | 0-72 | ⊕ | ⊕ | ■ | ■ | ■ | ■ | Fair |
| 30 | (Sobkoviak, Yount, & Halim, 2012) | Liberia | DHS 2006-2007 | 2467 | 0-48 | ■ | ⊕ | ■ | ⊕ | ⊕ | ⊕ | Good |
| 69 | (Subramanian, Ackerson, & Smith, 2010) | India | NFHS-3 2005-2006, and NFHS-4 2015-2016 | 19879 | 0-59 | ■ | ⊕ | ■ | ⊕ | ■ | ■ | Good |
| 87 | (Subramanyam, Kawachi, Berkman, & Subramanian, 2010) | India | NFHS 1992, 1998 and 2005 | 78310 | 0-35 | ⊕ | ⊕ | ■ | | ■ | ■ | Good |
| 35 | (Sunil, 2009) | Yemen | DHS 1997 | 10414 | 0-59 | ■ | ⊕ | ■ | ⊕ | ■ | ■ | Good |
| 41 | (Tariq, Sajjad, Zakar, Zakar, & Fischer, 2018) | Pakistan | PDHS 2012-2013 | 984 | 0-23 | ■ | ⊕ | ■ | ⊕ | ■ | ■ | Good |
| 59 | (Tesfaw & Dessie, 2022) | Ethiopia | EMDHS 2019 | 5027 | 0-59 | ■ | ⊕ | ■ | ⊕ | ■ | ■ | Good |
| 61 | (Tesfaw & Fenta, 2021) | Nigeria | DHS 2018 | 11314 | 0-59 | ■ | ⊕ | ■ | ⊕ | ■ | ■ | Good |

| | | | | | | | | | | | | |
|---|---|---|---|---|---|---|---|---|---|---|---|---|
| 92 | (Tibebu, Emiru, Tiruneh, Getu, & Azanaw, 2020) | Ethiopia | Cross-sectional data | 634 | 6 - 59 | ⊕ | ⊕ | ■ | ⊕ | ⊕ | ■ | Good |
| 71 | (Tiwari, Acharya, Paudel, Sapkota, & Kafle, 2020) | Nepal | DHS 2016 | 2355 | 0-59 | ⊕ | ⊕ | ■ | ⊕ | ■ | ■ | Good |
| 75 | (Li et al., 2022) | China | Cross-sectional data | 5529 | 0-71 | ■ | ⊕ | ■ | ⊕ | ■ | ■ | Good |
| **Cohort studies** | | | | | | | | | | | | |
| 34 | (Dadi, Miller, Woodman, Azale, & Mwanri, 2021) | Ethiopia | Study data | 866 | 0-72 | ⊕ | ■ | ⊕ | ⊕ | ⊕ | ■ | Fair |
| 58 | (R. Das et al., 2022) | Bangladesh | ABCD trial | 1431 | 0-23 | ■ | ⊕ | ■ | ⊕ | ■ | ■ | Good |
| 11 | (Fall et al., 2015) | Brazil, Guatemala, India, Philippines, and South Africa | COHORTs | 19403 | 24** | ■ | ⊕ | ⊕ | ⊕ | ■ | ⊕ | Fair |
| 54 | (Le Roux et al., 2019) | South Africa | Cross-sectional data | 470 | 0-24*** | ■ | ⊕ | ■ | ⊕ | ■ | ■ | Fair |
| 3 | (Schott et al., 2017) | Ethiopia, India | Young lives study | 343 | 0-78 | ⊕ | ⊕ | ■ | ⊕ | ■ | ⊕ | Fair |
| **Case control studies** | | | | | | | | | | | | |
| 74 | (Abitew, Yalew, Bezabih, & Bazzano, 2020) | Ethiopia | Baseline data: community-based programme records | 1273 | 6-59 | ■ | ⊕ | ⊕ | ⊕ | ■ | ■ | Fair |
| 24 | (Gebremaryam, Amare, Ayalew, Tigabu, & Menshaw, 2022) | Ethiopia | Bahir Dar City public hospitals | 67 cases, 134 controls | 6-23 | ⊕ | ■ | ⊕ | ⊕ | ⊕ | ■ | Good |
| 43 | (Owoaje, Onifade, & Desmennu, 2014) | Nigeria | Oni Memorial Hospital data | 100 cases, 200 controls | 6-23 | ⊕ | ⊕ | ■ | ⊕ | ⊕ | ■ | Fair |
| 25 | (Pravana et al., 2017) | Nepal | Community based random selection | 146 cases, 146 controls | 0-59 | ⊕ | ■ | ⊕ | ⊕ | ■ | ■ | Fair |
| 89 | (Wemakor, Azongo, Garti, & Atosona, 2018) | Ghana | Study questionnaire | 150 cases, 150 controls | 6-59 | ■ | ⊕ | ■ | ⊕ | ■ | ■ | Fair |
| **Randomised controlled trials** | | | | | | | | | | | | |
| 4 | (Friebert, Callaghan-Gillespie, Papathakis, & Manary, 2017) | Malawi | Mamachiponde study | 2284 | 6 weeks and 12 weeks** | ■ | ⊕ | ■ | ⊕ | ■ | ⊕ | Good |

■ Yes
⊕ No
-- other studies cited

*disaggregated by maternal age

**cohort study: measurements taken at specific time point

***longitudinal cohort study, measurements taken at 3, 6, 9, 12 and 24 months

[a] Moderate wasting: weight-for-height z-score (WHZ) <-2, weight-for-length z-score (WLZ) <-2, mid-upper arm circumference (MUAC) <125mm, and/or bilateral oedema

[b] Severe wasting: weight-for-height z-score (WHZ) <-3, weight-for-length z-score (WLZ) <-3, mid-upper arm circumference (MUAC) ≤115mm, and/or bilateral oedema

[c] Moderate underweight: weight-for-age z-score (WAZ) <-2

[d] Severe underweight: weight-for-age z-score (WAZ) <-3

**Supplementary Appendix 3. Reference list for all studies included in the systematic review and meta-analysis**


Abitew, D. B., Yalew, A. W., Bezabih, A. M., & Bazzano, A. N. (2020). Predictors of relapse of acute malnutrition following exit from community-based management program in Amhara region, Northwest Ethiopia: An unmatched case-control study. *PLoS ONE [Electronic Resource], 15*(4), e0231524. doi:https://dx.doi.org/10.1371/journal.pone.0231524

Aboagye, R. G., Seidu, A. A., Ahinkorah, B. O., Arthur-Holmes, F., Cadri, A., Dadzie, L. K., . . . Yaya, S. (2021). Dietary diversity and undernutrition in children aged 6-23 months in sub-Saharan Africa. *Nutrients, 13*(10). doi:http://dx.doi.org/10.3390/nu13103431

Adhikari, T., Yadav, J., Tripathi, N., Tolani, H., Kaur, H., & Rao, M. V. V. (2021). Do tribal children experience elevated risk of poor nutritional status in India? A multilevel analysis. *Journal of Biosocial Science, 53*(5), 683-708. doi:https://dx.doi.org/10.1017/S0021932020000474

Ahmed, A. S., Ahmed, T., Roy, S. K., Alam, N., & Hossain, M. I. (2012). Determinants of undernutrition in children under 2 years of age from Rural Bangladesh. *Indian Pediatrics, 49(10)*, 821-824. doi:http://dx.doi.org/10.1007/s13312-012-0187-2

Akseer, N., Bhatti, Z., Mashal, T., Soofi, S., Moineddin, R., Black, R. E., & Bhutta, Z. A. (2018). Geospatial inequalities and determinants of nutritional status among women and children in Afghanistan: An observational study. *The Lancet Global Health, 6*(4), e447-e459. doi:https://dx.doi.org/10.1016/S2214-109X(18)30025-1

Ali, N. B., Tahsina, T., Emdadul Hoque, D. M., Hasan, M. M., Iqbal, A., Huda, T. M., & El Arifeen, S. (2019). Association of food security and other socioeconomic factors with dietary diversity and nutritional statuses of children aged 6-59 months in rural Bangladesh. *PLoS ONE [Electronic Resource], 14(8) (no pagination)*. doi:http://dx.doi.org/10.1371/journal.pone.0221929

Alkaff, F. F., Flynn, J., Sukmajaya, W. P., & Salamah, S. (2020). Comparison of WHO growth standard and national Indonesian growth reference in determining prevalence and determinants of stunting and underweight in children under five: A cross-sectional study from Musi sub-district. *F1000Research, 9*. doi:https://dx.doi.org/10.12688/f1000research.23156.2

Amadu, I., Seidu, A. A., Duku, E., Okyere, J., Hagan, J. E., Hormenu, T., & Ahinkorah, B. O. (2021). The joint effect of maternal marital status and type of household cooking fuel on child nutritional status in sub-Saharan Africa: Analysis of cross-sectional surveys on children from 31 countries. *Nutrients, 13*(5). doi:http://dx.doi.org/10.3390/nu13051541

Amare, Z. Y., Ahmed, M. E., & Mehari, A. B. (2019). Determinants of nutritional status among children under age 5 in Ethiopia: Further analysis of the 2016 Ethiopia Demographic and Health Survey. *Globalization and Health, 15*(1). doi:http://dx.doi.org/10.1186/s12992-019-0505-7

Beiersmann, C., Lorenzo, J. B., Bountogo, M., Tiendrebeogo, J., Gabrysch, S., Ye, M., . . . Mu ller, O. (2013). Malnutrition determinants in young children from Burkina Faso. *Journal of Tropical Pediatrics, 59(5)*, 372-379. doi:http://dx.doi.org/10.1093/tropej/fmt037

Bekele, T., Rawstorne, P., & Rahman, B. (2021). Socioeconomic inequalities in child growth failure in Ethiopia: Findings from the 2000 and 2016 Demographic and Health Surveys. *BMJ Open, 11*(12). doi:https://dx.doi.org/10.1136/bmjopen-2021-051304

Birhan, N. A., & Belay, D. B. (2021). Associated risk factors of underweight among under-five children in Ethiopia using multilevel ordinal logistic regression model. *African Health Sciences, 21(1)*, 362-372. doi:http://dx.doi.org/10.4314/ahs.v21i1.46

Biswas, T., Townsend, N., Magalhaes, R. J. S., Hasan, M., & Mamun, A. (2021). Patterns and determinants of the double burden of malnutrition at the household level in South and Southeast Asia. *European Journal of Clinical Nutrition, 75*(2), 385-391. doi:https://dx.doi.org/10.1038/s41430-020-00726-z

Boah, M., Azupogo, F., Amporfro, D. A., & Abada, L. A. (2019). The epidemiology of undernutrition and its determinants in children under five years in Ghana. *PLoS ONE, 14*(7), e0219665. doi:10.1371/journal.pone.0219665

Dadi, A. F., Miller, E. R., Woodman, R. J., Azale, T., & Mwanri, L. (2021). Effect of perinatal depression on risk of adverse infant health outcomes in mother-infant dyads in Gondar town: A causal analysis. *BMC Pregnancy & Childbirth, 21*(1), 255. doi:https://dx.doi.org/10.1186/s12884-021-03733-5

Das, R., Kabir, M. F., Ashorn, P., Simon, J., Chisti, M. J., & Ahmed, T. (2022). Maternal underweight and its association with composite index of anthropometric failure among children under two years of age with diarrhea in Bangladesh. *Nutrients, 14*(9). doi:https://dx.doi.org/10.3390/nu14091935

Das, S., Fahim, S. M., Islam, M. S., Biswas, T., Mahfuz, M., & Ahmed, T. (2019). Prevalence and sociodemographic determinants of household-level double burden of malnutrition in Bangladesh. *Public Health Nutrition, 22*(8), 1425-1432. doi:https://dx.doi.org/10.1017/S1368980018003580

Fagbamigbe, A. F., Kandala, N. B., & Uthman, O. A. (2020). Decomposing the educational inequalities in the factors associated with severe acute malnutrition among under-five children in low- and middle-income countries. *BMC Public Health, 20*(1), 555. doi:https://dx.doi.org/10.1186/s12889-020-08635-3

Faisal Ahmed, N. A. M., Sultana, M., Ali, M., Abedin, M. M., Ahammed, B., Yeasmin, M. A., & Maniruzzaman, M. (2021). Identifying the factors causing malnutrition and its impact on mortality among under-five bangladeshi children. *Family Medicine and Primary Care Review, 23(3)*, 255-260. doi:http://dx.doi.org/10.5114/fmpcr.2021.108185

Fall, C. H., Sachdev, H. S., Osmond, C., Restrepo-Mendez, M. C., Victora, C., Martorell, R., . . . investigators, C. (2015). Association between maternal age at childbirth and child and adult outcomes in the offspring: A prospective study in five low-income and middle-income countries (COHORTS collaboration). *The Lancet Global Health, 3*(7), e366-377. doi:https://dx.doi.org/10.1016/S2214-109X(15)00038-8

Finlay, J. E., Ozaltin, E., & Canning, D. (2011). The association of maternal age with infant mortality, child anthropometric failure, diarrhoea and anaemia for first births: Evidence from 55 low- and middle-income countries. *BMJ Open, 1*(2), e000226. doi:https://dx.doi.org/10.1136/bmjopen-2011-000226

Friebert, A., Callaghan-Gillespie, M., Papathakis, P. C., & Manary, M. J. (2017). Adolescent pregnancy and nutrition: A subgroup analysis from the Mamachiponde study in Malawi. *Annals of the New York Academy of Sciences, 16*, 16. doi:https://dx.doi.org/10.1111/nyas.13465

Fuada, N., Latifah, L., Yunitawat, D., & Ashar, H. (2020). Assessment of nutritional status of children under-five in families of adolescent mothers in Indonesia 2013. *Journal of Nutritional Science & Vitaminology, 66*(Supplement), S425-S431. doi:https://dx.doi.org/10.3177/jnsv.66.S425

Gbadamosi, M. A., Goon, D. T., & Tugli, A. (2017). Relationship between feeding practices and patterns of infant growth: A cross-sectional study. *Research Journal of Medical Sciences, 11(4)*, 166-173. doi:http://dx.doi.org/10.3923/rjmsci.2017.166.173

Gebremaryam, T., Amare, D., Ayalew, T., Tigabu, A., & Menshaw, T. (2022). Determinants of severe acute malnutrition among children aged 6-23 months in Bahir Dar City public hospitals, Northwest Ethiopia, 2020: A case control study. *BMC Pediatrics, 22*(1). doi:https://dx.doi.org/10.1186/s12887-022-03327-w

Geda, N. R., Feng, C. X., Henry, C. J., Lepnurm, R., Janzen, B., & Whiting, S. J. (2021). Multiple anthropometric and nutritional deficiencies in young children in Ethiopia: a multi-level analysis based on a nationally representative data. *BMC Pediatrics, 21*(1), 11. doi:https://dx.doi.org/10.1186/s12887-020-02467-1

Gewa, C. A., & Yandell, N. (2012). Undernutrition among Kenyan children: Contribution of child, maternal and household factors. *Public Health Nutrition, 15*(6), 1029-1038. doi:https://dx.doi.org/10.1017/S136898001100245X

Ghimire, U., Aryal, B. K., Gupta, A. K., & Sapkota, S. (2020). Severe acute malnutrition and its associated factors among children under-five years: A facility-based cross-sectional study. *BMC Pediatrics, 20*(1), 249. doi:https://dx.doi.org/10.1186/s12887-020-02154-1

Haque, R., Alam, K., Rahman, S. M., Mustafa, M. U. R., Ahammed, B., Ahmad, K., . . . Keramat, S. A. (2022). Nexus between maternal underweight and child anthropometric status in South and South-East Asian countries. *Nutrition, 98*. doi:https://dx.doi.org/10.1016/j.nut.2022.111628

Hien, N. N., & Hoa, N. N. (2009). Nutritional status and determinants of malnutrition in children under three years of age in Nghean, Vietnam. *Pakistan Journal of Nutrition, 8(7)*, 958-964. doi:http://dx.doi.org/10.3923/pjn.2009.958.964

Hien, N. N., & Kam, S. (2008). Nutritional status and the characteristics related to malnutrition in children under five years of age in Nghean, Vietnam. *Journal of Preventive Medicine & Public Health / Yebang Uihakhoe Chi, 41*(4), 232-240. doi:https://dx.doi.org/10.3961/jpmph.2008.41.4.232

Hiruy, A. F., Xiong, Q., Jin, Q., Zhao, J., Lin, X., He, S., . . . Ying, C. (2021). The association of feeding practices and sociodemographic factors on underweight and wasting in children in Ethiopia: A secondary analysis of four health surveys from 2000 to 2016. *Journal of Tropical Pediatrics, 67*(4), 27. doi:https://dx.doi.org/10.1093/tropej/fmab047

Horta, B. L., Santos, R. V., Welch, J. R., Cardoso, A. M., Dos Santos, J. V., Assis, A. M. O., . . . Coimbra Jr, C. E. A. (2013). Nutritional status of indigenous children: Findings from the First National Survey of Indigenous People's Health and Nutrition in Brazil. *International Journal for Equity in Health, 12*(1). doi:http://dx.doi.org/10.1186/1475-9276-12-23

Hossain, F. B., Shawon, M. S. R., Al-Abid, M. S. U., Mahmood, S., Adhikary, G., & Bulbul, M. M. I. (2020). Double burden of malnutrition in children aged 24 to 59 months by socioeconomic status in five South Asian countries: Evidence from demographic and health surveys. *BMJ Open, 10(3) (no pagination)*. doi:http://dx.doi.org/10.1136/bmjopen-2019-032866

Hossain, M. B., & Khan, M. H. R. (2018). Role of parental education in reduction of prevalence of childhood undernutrition in Bangladesh. *Public Health Nutrition, 21*(10), 1845-1854. doi:https://dx.doi.org/10.1017/S1368980018000162

Huynh, G., Huynh, Q. H. N., Nguyen, N. H. T., Do, Q. T., & Khanh Tran, V. (2019). Malnutrition among 6-59-month-old children at District 2 Hospital, Ho Chi Minh

City, Vietnam: Prevalence and associated factors. *BioMed Research International, 2019*, 6921312. doi:http://dx.doi.org/10.1155/2019/6921312
Ickes, S. B., Hurst, T. E., & Flax, V. L. (2015). Maternal literacy, facility birth, and education are positively associated with better infant and young child feeding practices and nutritional status among Ugandan children. *Journal of Nutrition, 145(11)*, 2578-2586. doi:http://dx.doi.org/10.3945/jn.115.214346
Islam, M. S., & Biswas, T. (2020). Prevalence and correlates of the composite index of anthropometric failure among children under 5 years old in Bangladesh. *Maternal & Child Nutrition*, *16*(2), e12930. doi:https://dx.doi.org/10.1111/mcn.12930
Issah, A. N., Yeboah, D., Kpordoxah, M. R., Boah, M., & Mahama, A. B. (2022). Association between exposure to intimate partner violence and the nutritional status of women and children in Nigeria. *PLoS ONE [Electronic Resource]*, *17*(5), e0268462. doi:https://dx.doi.org/10.1371/journal.pone.0268462
Janevic, T., Petrovic, O., Bjelic, I., & Kubera, A. (2010). Risk factors for childhood malnutrition in Roma settlements in Serbia. *BMC Public Health*, *10*, 509. doi:https://dx.doi.org/10.1186/1471-2458-10-509
Karim, M. R., Al Mamun, A. S. M., Rana, M. M., Mahumud, R. A., Shoma, N. N., Dutt, D., . . . Hossain, M. G. (2021). Acute malnutrition and its determinants of preschool children in Bangladesh: Gender differentiation. *BMC Pediatrics*, *21*(1), 573. doi:https://dx.doi.org/10.1186/s12887-021-03033-z
Kasaye, H. K., Bobo, F. T., Yilma, M. T., & Woldie, M. (2019). Poor nutrition for under-five children from poor households in Ethiopia: Evidence from 2016 Demographic and Health Survey. *PLoS ONE*, *14*(12), e0225996. doi:10.1371/journal.pone.0225996
Khan, M. S., Halder, H. R., Rashid, M., Afroja, S., & Islam, M. (2020). Impact of socioeconomic and demographic factors for underweight and overweight children in Bangladesh: A polytomous logistic regression model. *Clinical Epidemiology and Global Health*, *8(4)*, 1348-1355. doi:http://dx.doi.org/10.1016/j.cegh.2020.05.010
Khan, S., Zaheer, S., & Safdar, N. F. (2019). Determinants of stunting, underweight and wasting among children <5 years of age: Evidence from 2012-2013 Pakistan Demographic and Health Survey. *BMC Public Health*, *19*(1), 358. doi:https://dx.doi.org/10.1186/s12889-019-6688-2
Kim, R., Rajpal, S., Joe, W., Corsi, D. J., Sankar, R., Kumar, A., & Subramanian, S. V. (2019). Assessing associational strength of 23 correlates of child anthropometric failure: An econometric analysis of the 2015-2016 National Family Health Survey, India. *Social Science & Medicine*, *238*, 112374. doi:https://dx.doi.org/10.1016/j.socscimed.2019.112374
Kumar, A., & Ram, F. (2013). Influence of family structure on child health: Evidence from India. *Journal of Biosocial Science*, *45*(5), 577-599. doi:https://dx.doi.org/10.1017/S0021932012000764
Kumar, R., Abbas, F., Mahmood, T., & Somrongthong, R. (2019). Prevalence and factors associated with underweight children: A population-based subnational analysis from Pakistan. *BMJ Open*, *9*(7). doi:http://dx.doi.org/10.1136/bmjopen-2019-028972
Kumar, R., & Paswan, B. (2021). Changes in socio-economic inequality in nutritional status among children in EAG states, India. *Public Health Nutrition*, *24*(6), 1304-1317. doi:https://dx.doi.org/10.1017/S1368980021000343
Le Roux, K., Christodoulou, J., Stansert-Katzen, L., Dippenaar, E., Laurenzi, C., Le Roux, I. M., . . . Rotheram-Borus, M. J. (2019). A longitudinal cohort study of rural adolescent vs adult South African mothers and their children from birth to 24 months. *BMC Pregnancy and Childbirth*, *19*(1). doi:http://dx.doi.org/10.1186/s12884-018-2164-8

Li, H., Yuan, S., Fang, H., Huang, G., Huang, Q., Wang, H., & Wang, A. (2022). Prevalence and associated factors for stunting, underweight and wasting among children under 6 years of age in rural Hunan Province, China: A community-based cross-sectional study. *BMC Public Health, 22*(1), 483. doi:https://dx.doi.org/10.1186/s12889-022-12875-w

Linnemayr, S., Alderman, H., & Ka, A. (2008). Determinants of malnutrition in Senegal: Individual, household, community variables, and their interaction. *Economics & Human Biology, 6*(2), 252-263. doi:https://dx.doi.org/10.1016/j.ehb.2008.04.003

Mashal, T., Takano, T., Nakamura, K., Kizuki, M., Hemat, S., Watanabe, M., & Seino, K. (2008). Factors associated with the health and nutritional status of children under 5 years of age in Afghanistan: Family behaviour related to women and past experience of war-related hardships. *BMC Public Health, 8*. doi:http://dx.doi.org/10.1186/1471-2458-8-301

Masibo, P. K., Humwa, F., & Macharia, T. N. (2020). The double burden of overnutrition and undernutrition in mother-child dyads in Kenya: Demographic and Health Survey data, 2014. *Journal of Nutritional Science, 9*, e5. doi:https://dx.doi.org/10.1017/jns.2019.39

Mena-Melendez, L. (2020). Ethnoracial child health inequalities in Latin America: Multilevel evidence from Bolivia, Colombia, Guatemala, and Peru. *SSM - Population Health, 12*. doi:http://dx.doi.org/10.1016/j.ssmph.2020.100673

Mokwena, K., & Kachabe, J. (2022). Profile of mothers whose children are treated for malnutrition at a rural district hospital in the North West province, South Africa. *South African Journal of Clinical Nutrition, 35(1)*, 17-22. doi:https://dx.doi.org/10.1080/16070658.2021.1921899

Mutunga, M., Frison, S., Rava, M., & Bahwere, P. (2020). The forgotten agenda of wasting in Southeast Asia: Burden, determinants and overlap with stunting: A review of nationally representative cross-sectional Demographic and Health Surveys in six countries. *Nutrients, 12*(2). doi:http://dx.doi.org/10.3390/nu12020559

Nakamori, M., Ninh, N. X., Khan, N. C., Huong, C. T., Tuan, N. A., Mai, L. B., . . . Yamamoto, S. (2010). Nutritional status, feeding practice and incidence of infectious diseases among children aged 6 to 18 months in northern mountainous Vietnam. *Journal of Medical Investigation, 57(1-2)*, 45-53. doi:http://dx.doi.org/10.2152/jmi.57.45

Nguyen, P. H., Sanghvi, T., Tran, L. M., Afsana, K., Mahmud, Z., Aktar, B., . . . Menon, P. (2017). The nutrition and health risks faced by pregnant adolescents: Insights from a cross-sectional study in Bangladesh. *PLoS ONE, 12*(6), e0178878. doi:https://dx.doi.org/10.1371/journal.pone.0178878

Nguyen, P. H., Scott, S., Khuong, L. Q., Pramanik, P., Ahmed, A., Rashid, S. F., . . . Menon, P. (2021). Adolescent birth and child undernutrition: An analysis of Demographic and Health Surveys in Bangladesh, 1996-2017. *Annals of the New York Academy of Sciences, 1500*(1), 69-81. doi:https://dx.doi.org/10.1111/nyas.14608

Nguyen, P. H., Scott, S., Neupane, S., Tran, L. M., & Menon, P. (2019). Social, biological, and programmatic factors linking adolescent pregnancy and early childhood undernutrition: A path analysis of India's 2016 National Family and Health Survey. *The Lancet Child & Adolescent Health, 3*(7), 463-473. doi:https://dx.doi.org/10.1016/S2352-4642(19)30110-5

Ntenda, P. A. M. (2019). Association of low birth weight with undernutrition in preschool-aged children in Malawi. *Nutrition Journal, 18*(1). doi:http://dx.doi.org/10.1186/s12937-019-0477-8

Ntenda, P. A. M., & Chuang, Y. C. (2018). Analysis of individual-level and community-level effects on childhood undernutrition in Malawi. *Pediatrics & Neonatology, 59*(4), 380-389. doi:https://dx.doi.org/10.1016/j.pedneo.2017.11.019

Obayelu, O. A., & Adeleye, O. R. (2021). Explaining child nutritional status in rural Nigeria: Socioeconomic dimensions. *Journal of Hunger and Environmental Nutrition, 16*(6), 829-846. doi:http://dx.doi.org/10.1080/19320248.2020.1781011

Olodu, M. D., Adeyemi, A. G., Olowookere, S. A., & Esimai, O. A. (2019). Nutritional status of under-five children born to teenage mothers in an urban setting, south-western Nigeria. *BMC Research Notes, 12*(1), 116. doi:https://dx.doi.org/10.1186/s13104-019-4147-x

Owoaje, E., Onifade, O., & Desmennu, A. (2014). Family and socioeconomic risk factors for undernutrition among children aged 6 to 23 Months in Ibadan, Nigeria. *The Pan African medical journal, 17*, 161. doi:https://dx.doi.org/10.11604/pamj.2014.17.161.2389

Paul, P., Chouhan, P., & Zaveri, A. (2019). Impact of child marriage on nutritional status and anaemia of children under 5 years of age: Empirical evidence from India. *Public Health, 177*, 95-101. doi:https://dx.doi.org/10.1016/j.puhe.2019.08.008

Paul, P., & Saha, R. (2022). Is maternal autonomy associated with child nutritional status? Evidence from a cross-sectional study in India. *PLoS ONE, 17*(5), e0268126. doi:https://dx.doi.org/10.1371/journal.pone.0268126

Poda, G. G., Hsu, C. Y., & Chao, J. C. J. (2017). Factors associated with malnutrition among children <5 years old in Burkina Faso: Evidence from the Demographic and Health Surveys IV 2010. *International Journal for Quality in Health Care, 29(7)*, 901-908. doi:http://dx.doi.org/10.1093/intqhc/mzx129

Porwal, A., Agarwal, P. K., Ashraf, S., Acharya, R., Ramesh, S., Khan, N., . . . Sarna, A. (2021). Association of maternal height and body mass index with nutrition of children under 5 years of age in India: Evidence from Comprehensive National Nutrition Survey 2016-18. *Asia Pacific Journal of Clinical Nutrition, 30*(4), 675-686. doi:https://dx.doi.org/10.6133/apjcn.202112_30(4).0014

Pramod Singh, G. C., Nair, M., Grubesic, R. B., & Connell, F. A. (2009). Factors associated with underweight and stunting among children in rural terai of Eastern Nepal. *Asia-Pacific Journal of Public Health, 21(2)*, 144-152. doi:https://dx.doi.org/10.1177/1010539509332063

Pravana, N. K., Piryani, S., Chaurasiya, S. P., Kawan, R., Thapa, R. K., & Shrestha, S. (2017). Determinants of severe acute malnutrition among children under 5 years of age in Nepal: A community-based case-control study. *BMJ Open, 7*(8). doi:http://dx.doi.org/10.1136/bmjopen-2017-017084

Qu, P., Mi, B., Wang, D., Zhang, R., Yang, J., Liu, D., . . . Yan, H. (2017). Association between the Infant and Child Feeding Index (ICFI) and nutritional status of 6- to 35-month-old children in rural western China. *PLoS ONE, 12*(2). doi:http://dx.doi.org/10.1371/journal.pone.0171984

Rahman, A., & Hossain, M. M. (2022). Quantile regression approach to estimating prevalence and determinants of child malnutrition. *Journal of Public Health, 30*(2), 323-339. doi:https://dx.doi.org/10.1007/s10389-020-01277-0

Rahman, M. A., Halder, H. R., Rahman, M. S., & Parvez, M. (2021). Poverty and childhood malnutrition: Evidence-based on a nationally representative survey of Bangladesh. *PLoS ONE, 16*(8), e0256235. doi:https://dx.doi.org/10.1371/journal.pone.0256235

Rahman, M. M. (2015). Is unwanted birth associated with child malnutrition in Bangladesh? *International Perspectives on Sexual & Reproductive Health, 41*(2), 80-88. doi:https://dx.doi.org/10.1363/4108015

Rahman, M. S., Rahman, M. A., Maniruzzaman, M., & Howlader, M. H. (2020). Prevalence of undernutrition in Bangladeshi children. *Journal of Biosocial Science, 52*(4), 596-609. doi:https://dx.doi.org/10.1017/S0021932019000683

Raj, A., Saggurti, N., Winter, M., Labonte, A., Decker, M. R., Balaiah, D., & Silverman, J. G. (2010). The effect of maternal child marriage on morbidity and mortality of children under 5 in India: Cross sectional study of a nationally representative sample. *BMJ, 340*, b4258. doi:https://dx.doi.org/10.1136/bmj.b4258

Rodgers, J., Kim, R., & Subramanian, S. V. (2020). Explaining within- vs between-population variation in child anthropometry and hemoglobin measures in India: A multilevel analysis of the National Family Health Survey 2015-2016. *Journal of Epidemiology, 30*(11), 485-496. doi:https://dx.doi.org/10.2188/jea.JE20190064

Samuel, A., Osendarp, S. J. M., Feskens, E. J. M., Lelisa, A., Adish, A., Kebede, A., & Brouwer, I. D. (2022). Gender differences in nutritional status and determinants among infants (6-11 m): A cross-sectional study in two regions in Ethiopia. *BMC public health, 22*(1), 401. doi:https://dx.doi.org/10.1186/s12889-022-12772-2

Sanjay, C., Gaurav, M., Suman, B., & Ramchandra, C. (2020). Determinants of nutritional status in children aged 0 - 6 years in rural Rajasthan. *Journal of Evolution of Medical and Dental Sciences, 9*(28), 1977-1983. doi:http://dx.doi.org/10.14260/jemds/2020/431

Schott, W., Aurino, E., Penny, M. E., & Behrman, J. R. (2017). Adolescent mothers' anthropometrics and grandmothers' schooling predict infant anthropometrics in Ethiopia, India, Peru, and Vietnam. *Annals of the New York Academy of Sciences, 24*, 24. doi:https://dx.doi.org/10.1111/nyas.13455

Sobkoviak, R. M., Yount, K. M., & Halim, N. (2012). Domestic violence and child nutrition in Liberia. *Social Science & Medicine, 74*(2), 103-111. doi:https://dx.doi.org/10.1016/j.socscimed.2011.10.024

Subramanian, S. V., Ackerson, L. K., & Smith, G. D. (2010). Parental BMI and childhood undernutrition in India: An assessment of intrauterine influence. *Pediatrics, 126*(3), e663-671. doi:https://dx.doi.org/10.1542/peds.2010-0222

Subramanyam, M. A., Kawachi, I., Berkman, L. F., & Subramanian, S. V. (2010). Socioeconomic inequalities in childhood undernutrition in India: Analyzing trends between 1992 and 2005. *PLoS ONE, 5*(6). doi:http://dx.doi.org/10.1371/journal.pone.0011392

Sunil, T. S. (2009). Effects of socio-economic and behavioural factors on childhood malnutrition in Yemen. *Maternal & Child Nutrition, 5*(3), 251-259. doi:https://dx.doi.org/10.1111/j.1740-8709.2008.00174.x

Tariq, J., Sajjad, A., Zakar, R., Zakar, M. Z., & Fischer, F. (2018). Factors associated with undernutrition in children under the age of two years: Secondary data analysis based on the Pakistan Demographic and Health Survey 2012-2013. *Nutrients, 10*(6). doi:http://dx.doi.org/10.3390/nu10060676

Tesfaw, L. M., & Dessie, Z. G. (2022). Multilevel multivariate analysis on the anthropometric indicators of under-five children in Ethiopia: EMDHS 2019. *BMC Pediatrics, 22*(1). doi:https://dx.doi.org/10.1186/s12887-022-03172-x

Tesfaw, L. M., & Fenta, H. M. (2021). Multivariate logistic regression analysis on the association between anthropometric indicators of under-five children in Nigeria: NDHS 2018. *BMC Pediatrics, 21*(1), 193. doi:https://dx.doi.org/10.1186/s12887-021-02657-5

Tibebu, N. S., Emiru, T. D., Tiruneh, C. M., Getu, B. D., & Azanaw, K. A. (2020). Underweight and its associated factors among children 6-59 months of age in Debre

Tabor Town, Amhara Region of Ethiopia, 2019: A community-based cross-sectional study. *Pediatric Health Med Ther*, *11*, 469-476. doi:10.2147/phmt.S288071

Tiwari, I., Acharya, K., Paudel, Y. R., Sapkota, B. P., & Kafle, R. B. (2020). Planning of births and childhood undernutrition in Nepal: evidence from a 2016 national survey. *BMC Public Health*, *20*(1), 1788. doi:https://dx.doi.org/10.1186/s12889-020-09915-8

Wemakor, A., Azongo, T., Garti, H., & Atosona, A. (2018). Young maternal age is a risk factor for child undernutrition in Tamale Metropolis, Ghana. *BMC research notes, 11(1)*, 877. doi:http://dx.doi.org/10.1186/s13104-018-3980-7

### Supplementary Appendix 4. Risk of bias and quality assessments

#### a. Cross-sectional and cohort studies

| ID | Year of Publication | Data Source | Questions | | | | | | | | | | | | | | Overall Rating |
|---|---|---|---|---|---|---|---|---|---|---|---|---|---|---|---|---|---|
| | | | 1 | 2 | 3 | 4 | 5 | 6 | 7 | 8 | 9 | 10 | 11 | 12 | 13 | 14 | |
| **Cross-sectional studies** | | | | | | | | | | | | | | | | | |
| 1 | 2021 | Cross-sectional study data | ■ | ■ | ■ | ■ | ■ | ⊕ | ⊕ | ■ | ■ | N/A | ■ | ⊕ | N/A | ■ | Good |
| 2 | 2021 | DHS Data 1996 - 2017 | ■ | ■ | ⊕ | ■ | ⊕ | ⊕ | ⊕ | ■ | ■ | N/A | ■ | ⊕ | N/A | ■ | Good |
| 5 | 2003 | India's NFHS-I data 1995 | ■ | ■ | ⊕ | ■ | ⊕ | ⊕ | ⊕ | ■ | ■ | N/A | ■ | ⊕ | N/A | ■ | Good |
| 6 | 2017 | Malawi DHS 2004 and 2010 | ■ | ■ | ⊕ | ■ | ⊕ | ⊕ | ⊕ | ■ | ■ | N/A | ■ | ⊕ | N/A | ■ | Good |
| 7 | 2019 | India NFHS - 4 2015 - 16 | ■ | ■ | ⊕ | ■ | ⊕ | ⊕ | ⊕ | ■ | ■ | N/A | ■ | ⊕ | N/A | ■ | Good |
| 8 | 2020 | Basic health research data 2013 (RISKESDAS 2013) | ■ | ■ | ⊕ | ■ | ⊕ | ⊕ | ⊕ | ■ | ■ | N/A | ⊕ | ⊕ | N/A | ⊕ | Fair |
| 9 | 2021 | Ethiopia DHS 2016 | ■ | ■ | ⊕ | ■ | ⊕ | ⊕ | ⊕ | ■ | ■ | N/A | ■ | ⊕ | N/A | ■ | Fair |
| 10 | 2022 | Nigeria DHS 2018 | ■ | ■ | ⊕ | ■ | ⊕ | ⊕ | ⊕ | ■ | ■ | N/A | ■ | ⊕ | N/A | ■ | Good |
| 12 | 2017 | Cross sectional survey conducted as part of the Rural Primary Health Care project | ■ | ■ | ⊕ | ■ | ⊕ | ⊕ | ⊕ | ■ | ■ | N/A | ■ | ⊕ | N/A | ■ | Good |
| 13 | 2021 | Ethiopia DHS 2000 - 2016 | ■ | ■ | ⊕ | ■ | ⊕ | ⊕ | ⊕ | ■ | ■ | N/A | ■ | ⊕ | N/A | ■ | Good |
| 14 | 2019 | Baseline data collected to assess the Nobokoli programme | ■ | ■ | ■ | ■ | ■ | ⊕ | ⊕ | ■ | ■ | ⊕ | ■ | ⊕ | N/A | ■ | Fair |
| 15 | 2019 | Malawi DHS 2015 -16 | ■ | ■ | ■ | ■ | ⊕ | ■ | ■ | ■ | ■ | ⊕ | ■ | ⊕ | N/A | ■ | Good |
| 16 | 2011 | 118 DHS surveys conducted in 55 countries between 1990 - 2008 | ■ | ■ | ■ | ■ | ⊕ | ⊕ | ⊕ | ■ | ■ | ⊕ | ■ | ⊕ | N/A | ■ | Good |
| 17 | 2021 | Comprehensive National Nutrition Survey (CNNS) India | ■ | ■ | ■ | ■ | ⊕ | ⊕ | ⊕ | ■ | ■ | ⊕ | ■ | ■ | N/A | ■ | Good |
| 18 | 2021 | India's NFHS-3 2005-2006, and NFHS-4 2015-2016 | ■ | ■ | ■ | ■ | ⊕ | ⊕ | ⊕ | ■ | ■ | ⊕ | ■ | ⊕ | N/A | ■ | Good |
| 19 | 2020 | Cross-sectional survey and secondary data from child health books and health record books | ■ | ■ | ⊕ | ■ | ⊕ | ⊕ | ⊕ | ■ | ■ | ⊕ | ■ | ⊕ | N/A | ■ | Good |
| 20 | 2020 | DHS survey data from 51 countries 2010 - 2018 | ■ | ⊕ | ■ | ■ | ⊕ | ⊕ | ⊕ | ■ | ■ | ⊕ | ■ | ⊕ | N/A | ■ | Good |
| 21 | 2008 | Baseline data collected for a nutrition intervention programme | ■ | ■ | ⊕ | ■ | ⊕ | ⊕ | ⊕ | ■ | ■ | ⊕ | ■ | ⊕ | N/A | ■ | Good |

| | | | | | | | | | | | | | | | | | |
|---|---|---|---|---|---|---|---|---|---|---|---|---|---|---|---|---|---|
| 22 | 2019 | Ethiopia DHS 2016 | ■ | ■ | ■ | ■ | ⊕ | ⊕ | ⊕ | ■ | ■ | ⊕ | ■ | ⊕ | N/A | ■ | Good |
| 23 | 2020 | Cross-sectional survey | ■ | ■ | ⊕ | ■ | ■ | ⊕ | ⊕ | ■ | ■ | ⊕ | ■ | ⊕ | N/A | ⊕ | Fair |
| 26 | 2019 | Pakistan DHS 2012 - 2013 | ■ | ■ | ■ | ■ | ⊕ | ⊕ | ⊕ | ■ | ■ | ⊕ | ■ | ⊕ | N/A | ■ | Good |
| 27 | 2012 | Bangladesh National Nutrition Programme baseline survey 2004 | ■ | ■ | ⊕ | ■ | ⊕ | ⊕ | ⊕ | ■ | ■ | ⊕ | ■ | ⊕ | N/A | ⊕ | Fair |
| 28 | 2021 | DHS survey data from 32 countries of Sub-Saharan Africa 2010 - 2020 | ■ | ■ | ■ | ■ | ⊕ | ⊕ | ⊕ | ■ | ■ | ⊕ | ■ | ⊕ | N/A | ■ | Good |
| 29 | 2021 | India NFHS - 4 2015 - 16 | ■ | ■ | ■ | ■ | ⊕ | ⊕ | ⊕ | ■ | ■ | ⊕ | ■ | ⊕ | N/A | ■ | Good |
| 30 | 2012 | Liberia DHS survey 2006-2007 | ■ | ■ | ■ | ■ | ⊕ | ⊕ | ⊕ | ■ | ■ | ⊕ | ■ | ⊕ | N/A | ■ | Good |
| 31 | 2020 | DHS surveys from Bangladesh, India, Pakistan, Maldives and Nepal between 2009 - 2016 | ■ | ■ | ■ | ■ | ⊕ | ⊕ | ⊕ | ■ | ■ | ⊕ | ■ | ■ | N/A | ■ | Good |
| 32 | 2020 | Kenya DHS survey 2014 | ■ | ■ | ■ | ■ | ⊕ | ⊕ | ⊕ | ■ | ■ | ⊕ | ■ | ⊕ | N/A | ■ | Good |
| 33 | 2010 | India's National Family Health Survey 2005 - 2006 | ■ | ■ | ■ | ■ | ⊕ | ⊕ | ⊕ | ■ | ■ | ⊕ | ■ | ■ | N/A | ■ | Good |
| 35 | 2009 | Yemen DHS survey 1997 | ■ | ■ | ⊕ | ■ | ⊕ | ⊕ | ⊕ | ■ | ■ | ⊕ | ■ | ■ | N/A | ■ | Good |
| 36 | 2019 | Ghana DHS 2014 | ■ | ■ | ■ | ■ | ⊕ | ⊕ | ⊕ | ■ | ■ | ⊕ | ■ | ⊕ | N/A | ■ | Good |
| 37 | 2020 | DHS data for Bolivia, Colombia, Guatemala and Peru 1986 - 2015 | ■ | ■ | ⊕ | ■ | ⊕ | ⊕ | ⊕ | ■ | ■ | ⊕ | ■ | ⊕ | N/A | ■ | Good |
| 38 | 2020 | India NFHS - 4 2015 - 16 | ■ | ■ | ⊕ | ■ | ⊕ | ⊕ | ⊕ | ■ | ■ | ⊕ | ■ | ⊕ | N/A | ■ | Good |
| 39 | 2017 | Burkino-Faso DHS 2010 | ■ | ■ | ⊕ | ■ | ⊕ | ⊕ | ⊕ | ■ | ■ | ⊕ | ■ | ⊕ | N/A | ■ | Good |
| 40 | 2008 | Cross-sectional survey data | ■ | ■ | ⊕ | ■ | ⊕ | ⊕ | ⊕ | ■ | ■ | ⊕ | ■ | ⊕ | N/A | ■ | Good |
| 41 | 2018 | PDHS 2012 - 2013 | ■ | ■ | ■ | ■ | ⊕ | ⊕ | ⊕ | ■ | ■ | ⊕ | ■ | ■ | N/A | ■ | Good |
| 42 | 2009 | Cross-sectional survey data | ■ | ■ | ⊕ | ■ | ■ | ⊕ | ⊕ | ■ | ■ | ⊕ | ■ | ⊕ | N/A | ■ | Good |
| 44 | 2020 | Cambodia DHS 2014, Lao PDR Multiple Indicator Survey (MICS) 2017, Myanmar DHS 2015, Thailand MICS 2015/16, Timor-Leste National Food and Nutrition Survey (NFNS) 2013, and Vietnam MICS 2011 | ■ | ■ | ⊕ | ■ | ⊕ | ⊕ | ⊕ | ■ | ■ | ⊕ | ■ | ⊕ | N/A | ■ | Good |
| 45 | 2022 | Baseline data collected for a large effectiveness study on the use of multi-micronutrient powders | ■ | ■ | ⊕ | ■ | ■ | ⊕ | ⊕ | ■ | ■ | ⊕ | ■ | ⊕ | N/A | ■ | Good |
| 46 | 2018 | ANNS 2013 (Afghanistan National Nutrition Survey) | ■ | ■ | ■ | ■ | ⊕ | ⊕ | ⊕ | ■ | ■ | ⊕ | ■ | ⊕ | N/A | ■ | Good |
| 47 | 2021 | Bangladesh DHS 2014 | ■ | ■ | ⊕ | ■ | ⊕ | ⊕ | ⊕ | ■ | ■ | ⊕ | ■ | ⊕ | N/A | ■ | Good |
| 48 | 2019 | India NFHS - 4 2015 - 16 | ■ | ■ | ⊕ | ■ | ⊕ | ⊕ | ⊕ | ■ | ■ | ⊕ | ■ | ⊕ | N/A | ■ | Good |
| 49 | 2020 | Bangladesh DHS 2014 | ■ | ■ | ⊕ | ■ | ⊕ | ⊕ | ⊕ | ■ | ■ | ⊕ | ■ | ■ | N/A | ■ | Good |

| | | | | | | | | | | | | | | | | |
|---|---|---|---|---|---|---|---|---|---|---|---|---|---|---|---|---|
| 50 | 2013 | India's NFHS 2005 - 2006 | ■ | ■ | ■ | ■ | ⊕ | ⊕ | ⊕ | ■ | ■ | ⊕ | ■ | ⊕ | N/A | ■ | Good |
| 51 | 2022 | India NFHS - 4 2015 - 16 | ■ | ■ | ■ | ■ | ⊕ | ⊕ | ⊕ | ■ | ■ | ⊕ | ■ | ⊕ | N/A | ■ | Good |
| 52 | 2015 | BDHS 2011 | ■ | ■ | ⊕ | ■ | ⊕ | ⊕ | ⊕ | ■ | ■ | ⊕ | ■ | ⊕ | N/A | ■ | Good |
| 53 | 2021 | DHS Data for 31 countries 2010 - 2019 | ■ | ⊕ | ⊕ | ■ | ⊕ | ⊕ | ⊕ | ■ | ■ | ⊕ | ■ | ⊕ | N/A | ■ | Good |
| 55 | 2019 | Cross-sectional study data | ■ | ■ | ⊕ | ■ | ⊕ | ⊕ | ⊕ | ■ | ■ | ⊕ | ■ | ⊕ | N/A | ■ | Good |
| 56 | 2013 | Cross-sectional study data | ■ | ■ | ⊕ | ■ | ■ | ⊕ | ⊕ | ■ | ■ | ⊕ | ■ | ⊕ | N/A | ■ | Good |
| 57 | 2015 | Uganda DHS 2006 and 2011 | ■ | ■ | ■ | ■ | ⊕ | ⊕ | ⊕ | ■ | ■ | ⊕ | ■ | ⊕ | N/A | ■ | Good |
| 59 | 2022 | EMDHS 2019 (Ethiopian mini DHS) | ■ | ■ | ⊕ | ■ | ⊕ | ⊕ | ⊕ | ■ | ■ | ⊕ | ■ | ⊕ | N/A | ■ | Good |
| 60 | 2021 | Ethiopia DHS 2016 | ■ | ■ | ■ | ■ | ⊕ | ⊕ | ⊕ | ■ | ■ | ⊕ | ■ | ⊕ | N/A | ■ | Good |
| 61 | 2021 | Nigeria DHS 2018 | ■ | ■ | ⊕ | ■ | ⊕ | ⊕ | ⊕ | ■ | ■ | ⊕ | ■ | ⊕ | N/A | ■ | Good |
| 62 | 2022 | DHS Data 2014 - 2018 | ■ | ■ | ■ | ■ | ⊕ | ⊕ | ⊕ | ■ | ■ | ⊕ | ■ | ⊕ | N/A | ■ | Good |
| 63 | 2017 | Baseline data from a scaled up MNCH programme | ■ | ■ | ⊕ | ■ | ⊕ | ⊕ | ⊕ | ■ | ■ | ⊕ | ■ | ⊕ | N/A | ■ | Good |
| 64 | 2009 | Cross-sectional study data | ■ | ■ | ⊕ | ■ | ■ | ⊕ | ⊕ | ■ | ■ | ⊕ | ■ | ⊕ | N/A | ■ | Good |
| 65 | 2008 | Cross-sectional study data | ■ | ■ | ⊕ | ■ | ■ | ⊕ | ⊕ | ■ | ■ | ⊕ | ■ | ⊕ | N/A | ■ | Good |
| 66 | 2013 | National Survey of Indigenous People's Health and Nutrition in Brazil 2008 - 2009 | ■ | ■ | ⊕ | ■ | ⊕ | ⊕ | ⊕ | ■ | ■ | ⊕ | ■ | ⊕ | N/A | ■ | Good |
| 67 | 2019 | Cross-sectional survey using primary health care centres records as the sampling frame | ■ | ■ | ■ | ■ | ■ | ⊕ | ⊕ | ■ | ■ | ⊕ | ■ | ⊕ | N/A | ■ | Good |
| 68 | 2010 | Cross-sectional study data | ■ | ■ | ■ | ■ | ⊕ | ⊕ | ⊕ | ■ | ■ | ⊕ | ■ | ⊕ | N/A | ⊕ | Fair |
| 69 | 2010 | India's NFHS-3 2005-2006, and NFHS-4 2015-2016 | ■ | ■ | ■ | ■ | ⊕ | ⊕ | ⊕ | ■ | ■ | ⊕ | ■ | ⊕ | N/A | ■ | Good |
| 70 | 2021 | DHS Survey data for Bangladesh, India, Nepal, Pakistan, Myanmar, Timor, Maldives and Cambodia conducted between 2007 and 2016 | ■ | ■ | ⊕ | ■ | ⊕ | ⊕ | ⊕ | ■ | ■ | ⊕ | ■ | ⊕ | N/A | ■ | Good |
| 71 | 2020 | Nepal DHS 2016 | ■ | ■ | ⊕ | ■ | ⊕ | ⊕ | ⊕ | ■ | ■ | ⊕ | ■ | ⊕ | N/A | ■ | Good |
| 72 | 2019 | Ethiopia DHS 2016 | ■ | ■ | ⊕ | ■ | ⊕ | ⊕ | ⊕ | ■ | ■ | ⊕ | ■ | ⊕ | N/A | ■ | Good |
| 73 | 2021 | Bangladesh DHS 2017-18 | ■ | ■ | ⊕ | ■ | ⊕ | ⊕ | ⊕ | ■ | ■ | ⊕ | ■ | ⊕ | N/A | ■ | Good |
| 75 | 2022 | Cross-sectional study data | ■ | ■ | ⊕ | ■ | ■ | ⊕ | ⊕ | ■ | ■ | ⊕ | ■ | ⊕ | N/A | ■ | Good |
| 76 | 2020 | Bangladesh DHS 2014 | ■ | ■ | ⊕ | ■ | ⊕ | ⊕ | ⊕ | ■ | ■ | ⊕ | ■ | ⊕ | N/A | ■ | Good |
| 77 | 2019 | MICS 2014 | ■ | ■ | ■ | ■ | ⊕ | ⊕ | ⊕ | ■ | ■ | ⊕ | ■ | ⊕ | N/A | ■ | Good |
| 78 | 2019 | Bangladesh DHS 2014 | ■ | ■ | ■ | ■ | ⊕ | ⊕ | ⊕ | ■ | ■ | ⊕ | ■ | ⊕ | N/A | ■ | Good |
| 79 | 2020 | Bangladesh DHS 2014 | ■ | ■ | ⊕ | ■ | ⊕ | ⊕ | ⊕ | ■ | ■ | ⊕ | ■ | ⊕ | N/A | ■ | Good |

| | | | | | | | | | | | | | | | | | |
|---|---|---|---|---|---|---|---|---|---|---|---|---|---|---|---|---|---|
| 80 | 2022 | Cross-sectional study data | ■ | ■ | ■ | ■ | ⊕ | ⊕ | ⊕ | ■ | ■ | ⊕ | ■ | ⊕ | N/A | ⊕ | Fair |
| 81 | 2017 | Cross-sectional data from the MAL-ED Cohort study | ■ | ■ | ■ | ■ | ⊕ | ⊕ | ⊕ | ■ | ■ | ⊕ | ■ | ⊕ | N/A | ■ | Good |
| 82 | 2010 | Serbia MICS 2005 | ■ | ■ | ■ | ■ | ⊕ | ⊕ | ⊕ | ■ | ■ | ⊕ | ■ | ⊕ | N/A | ■ | Good |
| 83 | 2018 | Bangladesh DHS 2014 | ■ | ■ | ⊕ | ■ | ⊕ | ⊕ | ⊕ | ■ | ■ | ⊕ | ■ | ⊕ | N/A | ■ | Good |
| 84 | 2020 | Cross-sectional study data from children admitted to the Outpatients Therapeutic Care Centers | ■ | ■ | ■ | ■ | ■ | ⊕ | ⊕ | ■ | ■ | ⊕ | ■ | ⊕ | N/A | ■ | Good |
| 85 | 2019 | India's NFHS - 4 2015 - 16 | ■ | ■ | ⊕ | ■ | ⊕ | ⊕ | ⊕ | ■ | ■ | ⊕ | ■ | ⊕ | N/A | ■ | Good |
| 86 | 2021 | Ethiopia DHS 2000 and 2016 | ■ | ■ | ⊕ | ■ | ⊕ | ⊕ | ⊕ | ■ | ■ | ⊕ | ■ | ⊕ | N/A | ■ | Good |
| 87 | 2010 | India's NFHS 1992, 1998 and 2005 | ■ | ■ | ■ | ■ | ⊕ | ⊕ | ⊕ | ■ | ■ | ⊕ | ■ | ⊕ | N/A | ■ | Good |
| 88 | 2012 | Kenya DHS survey 2003 | ■ | ■ | ■ | ■ | ⊕ | ⊕ | ⊕ | ■ | ■ | ⊕ | ■ | ⊕ | N/A | ■ | Good |
| 90 | 2021 | Nigeria DHS 2013 | ■ | ■ | ⊕ | ■ | ⊕ | ■ | ⊕ | ■ | ■ | ⊕ | ■ | ⊕ | N/A | ■ | Good |
| 91 | 2022 | Bangladesh DHS 2014 | ■ | ■ | ⊕ | ■ | ⊕ | ■ | ⊕ | ■ | ■ | ⊕ | ■ | ⊕ | N/A | ■ | Good |
| 92 | 2020 | Cross-sectional study data | ■ | ■ | ■ | ■ | ■ | ⊕ | ⊕ | ■ | ■ | ⊕ | ■ | ⊕ | N/A | ■ | Good |
| **Cohort Studies** | | | | | | | | | | | | | | | | | |
| 3 | 2017 | Secondary analysis of data from the Young lives study | ■ | ■ | ■ | ■ | ⊕ | ■ | ■ | ■ | ■ | ■ | ■ | ⊕ | N/A | ■ | Fair |
| 11 | 2015 | Data from COHORTs (Consortium for Health Orientated Research in Transitioning Societies) | ■ | ■ | ■ | ■ | ⊕ | ■ | ■ | ■ | ■ | ■ | ■ | ⊕ | ■ | ■ | Fair |
| 34 | 2021 | Study data | ■ | ■ | ⊕ | ■ | ■ | ⊕ | ⊕ | ■ | ■ | ⊕ | ■ | ⊕ | ■ | ■ | Fair |
| 54 | 2019 | Cross-sectional study data | ■ | ■ | ■ | ■ | ⊕ | ■ | ■ | ■ | ■ | ■ | ■ | ⊕ | ■ | ■ | Fair |
| 58 | 2022 | Data from ABCD trial Bangladesh Sit | ■ | ■ | ⊕ | ■ | ■ | ■ | ■ | ■ | ■ | ⊕ | ■ | ⊕ | ■ | ■ | Good |

■ = Yes
⊕ = No / not reported

**Questions:**

1. Was the research question or objective in this paper clearly stated?
2. Was the study population clearly specified and defined?
3. Was the participation rate of eligible persons at least 50%?
4. Were all the subjected selected or recruited from the same or similar populations (including the same time period)? Were inclusions and exclusion criteria for being in the study prespecified and applied uniformly to all participants?

5. Was a sample size justification, power description, or variance and effect estimates provided?
6. For the analyses in this paper, were the exposure(s) of interest measured prior to the outcome(s) being measured?
7. Was the timeframe sufficient so that one could reasonably expect to see an association between exposure and outcome if it existed?
8. For the exposures that can vary in amount or level, did the study examine different levels of the exposure as related to the outcomes e.g. categories of exposure or exposure measured as a continuous variable
9. Were the exposure measures (independent variables) clearly defined, valid, reliable and implemented consistently across all study participants?
10. Were the exposures measures assessed more than once over time?
11. Were the outcome measures clearly defined, valid, reliable and implemented consistently across all participants?
12. Were the outcome assessors blinded to the exposure status of the participants?
13. Was loss to follow-up after baseline 20% or less?
14. Were key potential confounding variables measured and adjusted statistically for their impact on the relationship between exposure(s) and outcome(s)?

### b. Case control studies

| ID | Year of Publication | Data Source | Questions 1 | 2 | 3 | 4 | 5 | 6 | 7 | 8 | 9 | 10 | 11 | 12 | Overall Rating |
|---|---|---|---|---|---|---|---|---|---|---|---|---|---|---|---|
| 24 | 2022 | Bahir Dar City public hospitals | ■ | ■ | ■ | ■ | ■ | ■ | ■ | ■ | ⊙ | ■ | ⊕ | ■ | Good |
| 25 | 2017 | Community based random selection | ■ | ■ | ■ | ■ | ■ | ■ | ⊕ | ■ | ■ | ■ | ⊕ | ⊙ | Fair |
| 43 | 2014 | Oni Memorial Hospital data | ■ | ■ | ■ | ■ | ■ | ■ | ⊕ | ⊕ | ⊙ | ⊕ | ⊕ | ⊙ | Fair |
| 74 | 2020 | Baseline data from community-based management programme records assessing factors associated with relapse | ■ | ■ | ■ | ■ | ■ | ■ | ⊕ | ⊕ | ■ | ■ | ⊕ | No | Fair |
| 89 | 2018 | Study questionnaire | ■ | ■ | ■ | ■ | ■ | ■ | ⊕ | ⊕ | ■ | ■ | ⊕ | ■ | Fair |

■ = Yes
⊕ = No / not reported
⊙ = Partial

**Questions:**

1. Was the research question or objective in this paper clearly stated?
2. Was the study population clearly specified and defined?
3. Did the authors include a sample size justification?
4. Were the controls selected or recruited from the same or similar population that gave rise to the cases (including the same timeframe)?
5. Were the definitions, inclusion and exclusion criteria, algorithms or processes used to identify or select cases and controls valid, reliable and implemented consistently across all study participants?
6. Were the cases clearly defined and differentiated from controls?
7. If less than 100 percent of eligible cases and/or controls were selected from the study, were the cases and/or controls randomly selected from those eligible?
8. Was there use of concurrent controls?
9. Were the investigators able to confirm that the exposure / risk occurred prior to the development of the condition or event that defined a participant as a case?
10. Were the measures of exposure/risk clearly defined, valid, reliable, and implemented consistently (including the same time period) across all study participants?
11. Were the assessors of exposure / risk blinded to the case or control status of the participants?
12. Were key potential confounding variables measured and adjusted statistically in the analyses? If matching was used, did the investigators account for matching during study analysis?

#### c. Randomised controlled trials

| ID | Year of Publication | Data Source | Questions | | | | | | | | | | | | | | Overall Rating |
|---|---|---|---|---|---|---|---|---|---|---|---|---|---|---|---|---|---|
| | | | 1 | 2 | 3 | 4 | 5 | 6 | 7 | 8 | 9 | 10 | 11 | 12 | 13 | 14 | |
| 4 | 2017 | Subgroup analysis from the Mamachiponde study | ■ | ■ | ■ | ⨁ | ■ | ■ | ■ | ■ | ■ | ■ | ■ | ■ | ⊙ | ■ | Good |

■ = Yes
⨁ = No / not reported
⊙ = Partial

**Questions:**

1. Was the study described as randomized, a randomized trial, a randomized clinical trial, or an RCT?
2. Was the method of randomization adequate (i.e., use of randomly generated assignment)?
3. Was the treatment allocation concealed (so that assignments could not be predicted)?
4. Were study participants and providers blinded to treatment group assignment?
5. Were the people assessing the outcomes blinded to the participants' group assignments?
6. Were the groups similar at baseline on important characteristics that could affect outcomes (e.g., demographics, risk factors, co-morbid conditions)?
7. Was the overall drop-out rate from the study at endpoint 20% or lower of the number allocated to treatment?
8. Was the differential drop-out rate (between treatment groups) at endpoint 15 percentage points or lower?
9. Was there high adherence to the intervention protocols for each treatment group?
10. Were other interventions avoided or similar in the groups (e.g., similar background treatments)?
11. Were outcomes assessed using valid and reliable measures, implemented consistently across all study participants?
12. Did the authors report that the sample size was sufficiently large to be able to detect a difference in the main outcome between groups with at least 80% power?
13. Were outcomes reported or subgroups analysed prespecified (i.e., identified before analyses were conducted)?
14. Were all randomized participants analysed in the group to which they were originally assigned, i.e., did they use an intention-to-treat analysis?

**Supplementary Figure 1. Association between adolescent pregnancy (10-19 years) and the pooled odds of moderate childhood wasting (1-59 months), versus adult pregnancy**

| Study | Treatment Yes | Treatment No | Control Yes | Control No | Odds ratio with 95% CI | Weight (%) |
|---|---|---|---|---|---|---|
| Karim et al | 9 | 32 | 38 | 461 | 3.41 [ 1.52, 7.67] | 1.01 |
| Rajaram et al | 29 | 179 | 268 | 1,773 | 1.07 [ 0.71, 1.62] | 2.29 |
| Kim et al | 10,413 | 40,014 | 17,058 | 72,959 | 1.11 [ 1.08, 1.14] | 4.12 |
| Ntenda | 11 | 228 | 68 | 2,165 | 1.54 [ 0.80, 2.95] | 1.38 |
| Finlay et al | 70,768 | 36,429 | 48,677 | 20,709 | 0.83 [ 0.81, 0.84] | 4.13 |
| Porwal et al | 57 | 322 | 5,255 | 25,095 | 0.85 [ 0.64, 1.12] | 3.00 |
| Amare et al | 89 | 904 | 851 | 7,576 | 0.88 [ 0.70, 1.10] | 3.31 |
| Khan et al | 141 | 1,140 | 177 | 1,613 | 1.13 [ 0.89, 1.42] | 3.29 |
| Aboagye et al | 447 | 3,981 | 5,424 | 39,117 | 0.81 [ 0.73, 0.90] | 3.94 |
| Raj et al | 1,754 | 8,478 | 739 | 4,263 | 1.19 [ 1.09, 1.31] | 3.97 |
| Sunil | 126 | 989 | 841 | 5,545 | 0.84 [ 0.69, 1.03] | 3.48 |
| Tariq et al | 5 | 156 | 39 | 784 | 0.64 [ 0.25, 1.66] | 0.79 |
| Mutunga et al | 185 | 1,622 | 3,180 | 37,321 | 1.34 [ 1.14, 1.57] | 3.71 |
| Faisal et al | 463 | 2,445 | 471 | 2,962 | 1.19 [ 1.04, 1.37] | 3.79 |
| Paul et al | 8,496 | 29,675 | 7,572 | 25,919 | 0.98 [ 0.95, 1.02] | 4.11 |
| Amadu et al | 886 | 8,344 | 9,874 | 110,542 | 1.19 [ 1.11, 1.28] | 4.04 |
| Le Roux et al | 11 | 65 | 37 | 421 | 1.93 [ 0.94, 3.96] | 1.19 |
| Le Roux et al | 5 | 71 | 18 | 440 | 1.72 [ 0.62, 4.78] | 0.70 |
| Le Roux et al | 2 | 74 | 8 | 450 | 1.52 [ 0.32, 7.30] | 0.33 |
| Le Roux et al | 4 | 72 | 14 | 444 | 1.76 [ 0.56, 5.50] | 0.58 |
| Le Roux et al | 4 | 72 | 14 | 444 | 1.76 [ 0.56, 5.50] | 0.58 |
| Das et al | 1 | 31 | 109 | 1,290 | 0.38 [ 0.05, 2.82] | 0.21 |
| Tesfaw and Dessie | 308 | 2,877 | 150 | 1,692 | 1.21 [ 0.98, 1.48] | 3.46 |
| Tesfaw and Fenta | 460 | 7,540 | 298 | 3,016 | 0.62 [ 0.53, 0.72] | 3.73 |
| Haque et al | 6,019 | 22,686 | 39,890 | 145,135 | 0.97 [ 0.94, 1.00] | 4.12 |
| Horta et al | 20 | 794 | 57 | 5,168 | 2.28 [ 1.36, 3.82] | 1.84 |
| Subramanian et al | 137 | 534 | 2,531 | 12,774 | 1.29 [ 1.07, 1.57] | 3.52 |
| Rahman et al | 321 | 3,366 | 357 | 3,692 | 0.99 [ 0.84, 1.15] | 3.70 |
| Abitew et al | 15 | 35 | 430 | 793 | 0.79 [ 0.43, 1.46] | 1.48 |
| Yuan et al | 2 | 23 | 219 | 5,285 | 2.10 [ 0.49, 8.96] | 0.38 |
| Islam and Biswas | 163 | 4,922 | 58 | 1,822 | 1.04 [ 0.77, 1.41] | 2.88 |
| Rahman et al | 1,192 | 4,014 | 275 | 1,650 | 1.78 [ 1.54, 2.06] | 3.77 |
| Janevic et al | 25 | 545 | 13 | 223 | 0.79 [ 0.40, 1.57] | 1.28 |
| Hossain and Khan | 181 | 869 | 849 | 5,273 | 1.29 [ 1.09, 1.54] | 3.61 |
| Nguyen et al | 3,081 | 10,159 | 9,534 | 33,506 | 1.07 [ 1.02, 1.12] | 4.09 |
| Wemakor et al | 18 | 132 | 6 | 144 | 3.27 [ 1.26, 8.49] | 0.78 |
| Obayelu and Adeleye | 220 | 1,895 | 1,600 | 13,484 | 0.98 [ 0.84, 1.14] | 3.74 |
| Rahman et al | 177 | 1,406 | 2,259 | 14,147 | 0.79 [ 0.67, 0.93] | 3.68 |
| **Overall** | | | | | 1.09 [ 0.99, 1.19] | |

Heterogeneity: $\tau^2 = 0.06$, $I^2 = 96.97\%$, $H^2 = 33.05$

Test of $\theta_i = \theta_j$: $Q(37) = 608.77$, $p = 0.00$

Test of $\theta = 0$: $z = 1.74$, $p = 0.08$

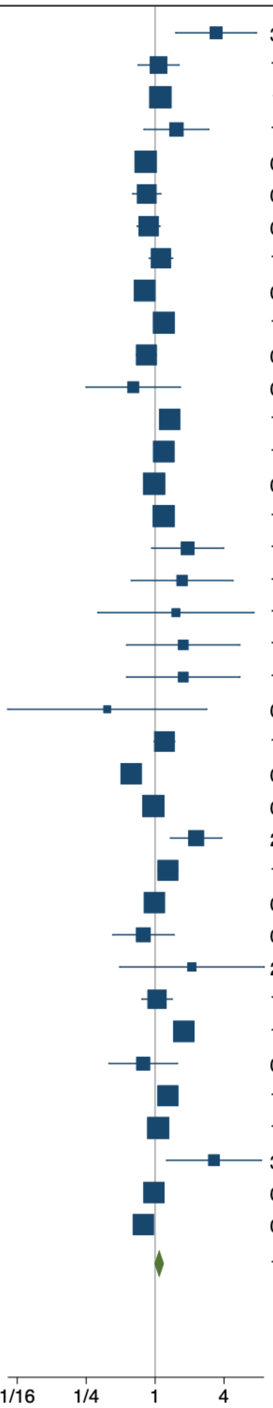


Random-effects REML model

Key: Treatment – number of children in adolescent mother group, control – number of children in adult mother group, yes – number of children with the outcome (moderate wasting, weight-for-height z-score (WHZ) or weight-for-length z-score (WLZ) <-2 to -3 or mid-upper arm circumference (MUAC) 125mm to >115mm), no – number of children without the outcome

**Supplementary Figure 2. Association between adolescent pregnancy (10-19 years) and the pooled odds of severe childhood wasting (1-59 months), versus adult pregnancy**

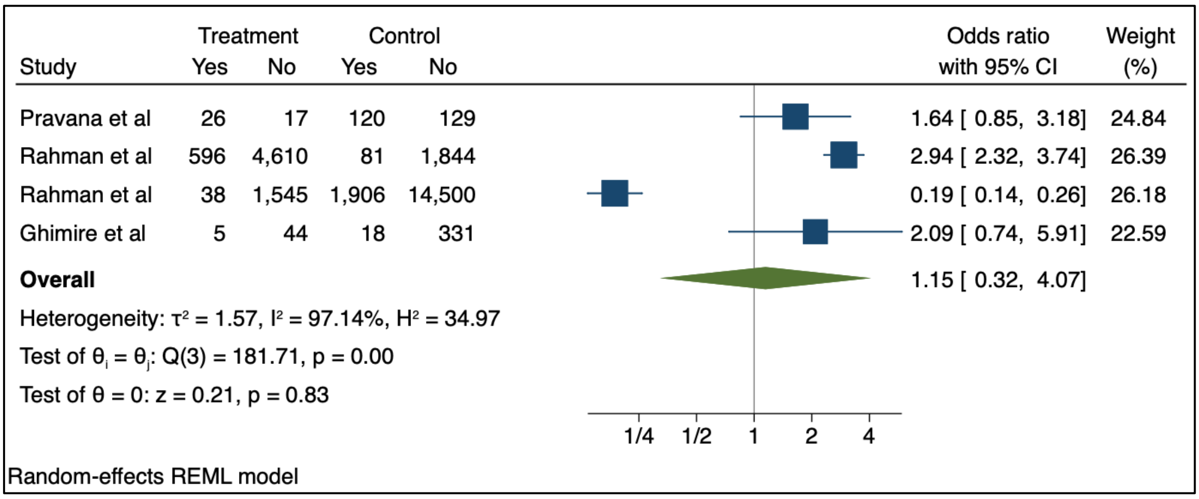


Key: Treatment – number of children in adolescent mother group, control – number of children in adult mother group, yes – number of children with the outcome (severe wasting, weight-for-height z-score (WHZ) or weight-for-length z-score (WLZ) <-3 or mid-upper arm circumference (MUAC) ≤115mm), no – number of children without the outcome

**Supplementary Figure 3. Association between adolescent pregnancy (10-19 years) and the pooled odds of moderate childhood underweight (1-59 months), versus adult pregnancy**

| Study | Treatment Yes | Treatment No | Control Yes | Control No | Odds ratio with 95% CI | Weight (%) |
|---|---|---|---|---|---|---|
| Rajaram et al | 76 | 132 | 490 | 1,551 | 1.82 [ 1.35, 2.46] | 2.80 |
| Kim et al | 20,216 | 30,211 | 27,797 | 62,220 | 1.50 [ 1.46, 1.53] | 2.96 |
| Birhan and Belay | 1,207 | 3,229 | 1,048 | 3,451 | 1.23 [ 1.12, 1.35] | 2.94 |
| Ntenda | 26 | 213 | 281 | 1,952 | 0.85 [ 0.55, 1.30] | 2.65 |
| Finlay et al | 72,150 | 35,047 | 49,647 | 19,739 | 0.82 [ 0.80, 0.84] | 2.96 |
| Porwal et al | 73 | 318 | 10,293 | 20,954 | 0.47 [ 0.36, 0.60] | 2.84 |
| Alkaff et al | 13 | 7 | 56 | 142 | 4.71 [ 1.79, 12.42] | 1.85 |
| Sanjay et al | 7 | 8 | 222 | 157 | 0.62 [ 0.22, 1.74] | 1.76 |
| Khan et al | 409 | 872 | 406 | 1,384 | 1.60 [ 1.36, 1.88] | 2.91 |
| Aboagye et al | 779 | 3,649 | 8,119 | 36,422 | 0.96 [ 0.88, 1.04] | 2.94 |
| Raj et al | 4,912 | 5,318 | 1,665 | 3,337 | 1.85 [ 1.73, 1.99] | 2.95 |
| Sunil | 527 | 588 | 2,931 | 3,455 | 1.06 [ 0.93, 1.20] | 2.93 |
| Tariq et al | 10 | 151 | 90 | 733 | 0.54 [ 0.27, 1.06] | 2.29 |
| Parmod et al | 122 | 87 | 114 | 120 | 1.48 [ 1.01, 2.15] | 2.71 |
| Faisal et al | 1,078 | 1,830 | 1,014 | 2,419 | 1.41 [ 1.27, 1.56] | 2.94 |
| Paul et al | 14,704 | 23,467 | 10,460 | 23,031 | 1.38 [ 1.34, 1.42] | 2.96 |
| Khan et al | 1,220 | 1,920 | 1,181 | 2,496 | 1.34 [ 1.22, 1.48] | 2.94 |
| Amadu et al | 1,632 | 7,598 | 20,868 | 99,548 | 1.02 [ 0.97, 1.08] | 2.95 |
| Le Roux et al | 7 | 69 | 14 | 444 | 3.22 [ 1.25, 8.25] | 1.89 |
| Le Roux et al | 4 | 72 | 18 | 440 | 1.36 [ 0.45, 4.13] | 1.66 |
| Le Roux et al | 2 | 74 | 14 | 444 | 0.86 [ 0.19, 3.85] | 1.22 |
| Le Roux et al | 2 | 74 | 18 | 440 | 0.66 [ 0.15, 2.91] | 1.23 |
| Le Roux et al | 2 | 74 | 14 | 444 | 0.86 [ 0.19, 3.85] | 1.22 |
| Tesfaw and Dessie | 792 | 2,393 | 380 | 1,462 | 1.27 [ 1.11, 1.46] | 2.92 |
| Tesfaw and Fenta | 1,582 | 6,418 | 842 | 2,472 | 0.72 [ 0.66, 0.80] | 2.94 |
| Haque et al | 11,208 | 17,497 | 63,315 | 121,710 | 1.23 [ 1.20, 1.26] | 2.96 |
| Horta et al | 42 | 772 | 314 | 4,911 | 0.85 [ 0.61, 1.18] | 2.76 |
| Subramanian et al | 260 | 411 | 5,321 | 9,984 | 1.19 [ 1.01, 1.39] | 2.91 |
| Tiwari et al | 99 | 198 | 539 | 1,517 | 1.41 [ 1.08, 1.83] | 2.83 |
| Rahman et al | 678 | 3,009 | 863 | 3,186 | 0.83 [ 0.74, 0.93] | 2.93 |
| Yuan et al | 3 | 22 | 214 | 5,290 | 3.37 [ 1.00, 11.35] | 1.53 |
| Islam and Biswas | 148 | 4,937 | 58 | 1,822 | 0.94 [ 0.69, 1.28] | 2.79 |
| Janevic et al | 39 | 531 | 15 | 221 | 1.08 [ 0.58, 2.00] | 2.38 |
| Hossain and Khan | 328 | 722 | 2,013 | 4,109 | 0.93 [ 0.81, 1.07] | 2.92 |
| Nguyen et al | 4,805 | 130,719 | 12,317 | 31,773 | 0.09 [ 0.09, 0.10] | 2.95 |
| Subramanyam et al | 3,341 | 3,964 | 28,332 | 42,673 | 1.27 [ 1.21, 1.33] | 2.95 |
| Wemakor et al | 44 | 106 | 5 | 145 | 12.04 [ 4.62, 31.39] | 1.87 |
| Obayelu and Adeleye | 491 | 1,624 | 6,056 | 9,028 | 0.45 [ 0.41, 0.50] | 2.94 |
| Rahman et al | 465 | 1,118 | 5,127 | 11,279 | 0.91 [ 0.82, 1.02] | 2.93 |
| **Overall** | | | | | 1.09 [ 0.87, 1.35] | |

Heterogeneity: $\tau^2 = 0.41$, $I^2 = 99.71\%$, $H^2 = 344.43$

Test of $\theta_i = \theta_j$: Q(38) = 19872.70, p = 0.00

Test of $\theta = 0$: z = 0.74, p = 0.46

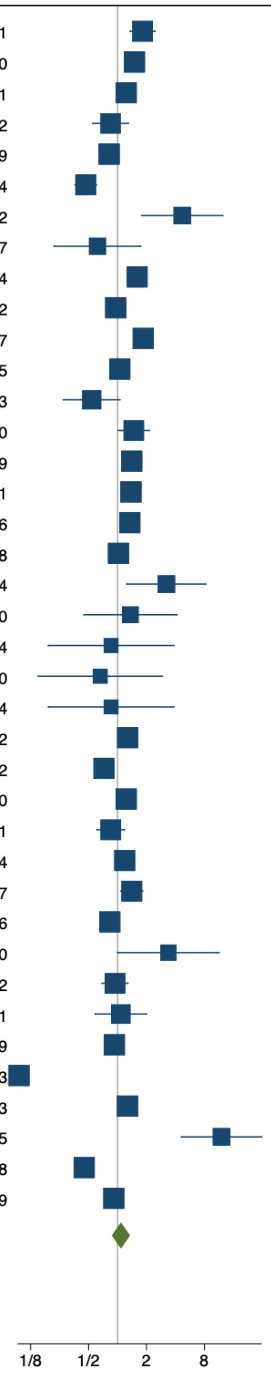


Random-effects REML model

Key: Treatment – number of children in adolescent mother group, control – number of children in adult mother group, yes – number of children with the outcome (moderate underweight, weight-for-age z-score (WAZ) <-2 to -3), no – number of children without the outcome

**Supplementary Figure 4. Association between adolescent pregnancy (10-24 years) and the pooled odds of moderate childhood wasting (1-59 months), versus adult pregnancy, stratified by region (Asia)**

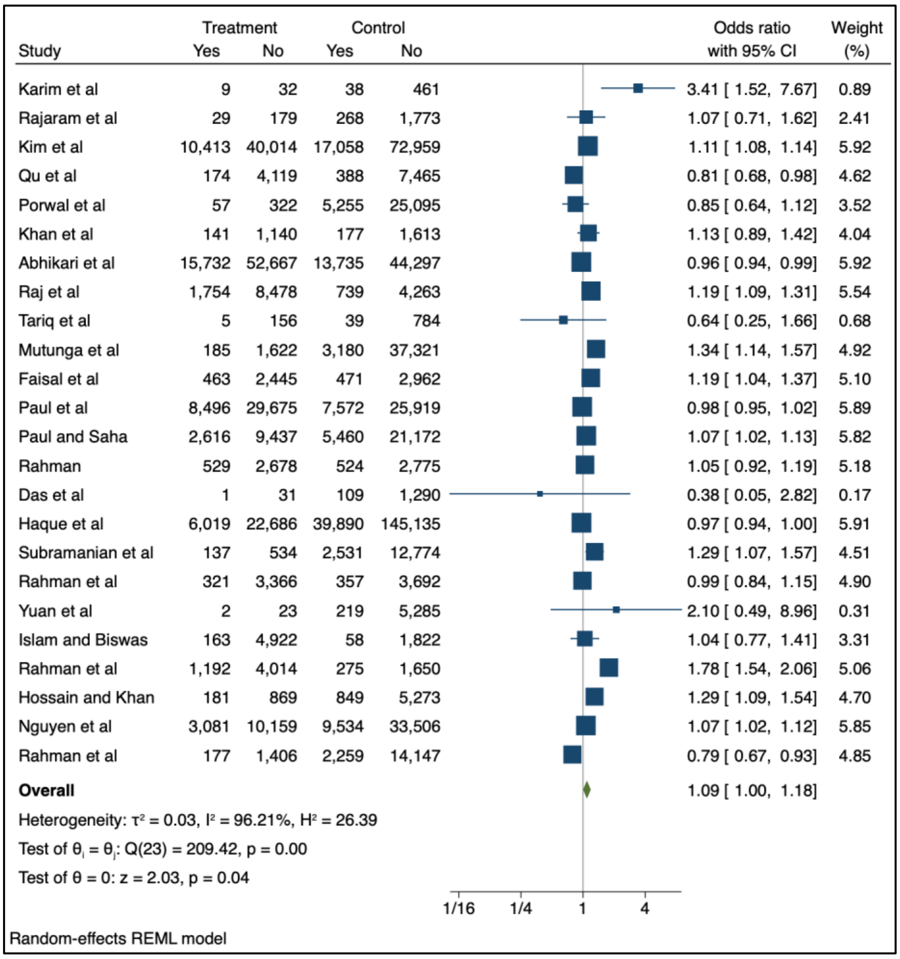


Key: Treatment – number of children in adolescent mother group, control – number of children in adult mother group, yes – number of children with the outcome (moderate wasting, weight-for-height z-score (WHZ) or weight-for-length z-score (WLZ) <-2 to -3 or mid-upper arm circumference (MUAC) 125mm to >115mm), no – number of children without the outcome

**Supplementary Figure 5. Association between adolescent pregnancy (10-24 years) and the pooled odds of moderate childhood underweight (1-59 months), versus adult pregnancy, stratified by region (Asia)**

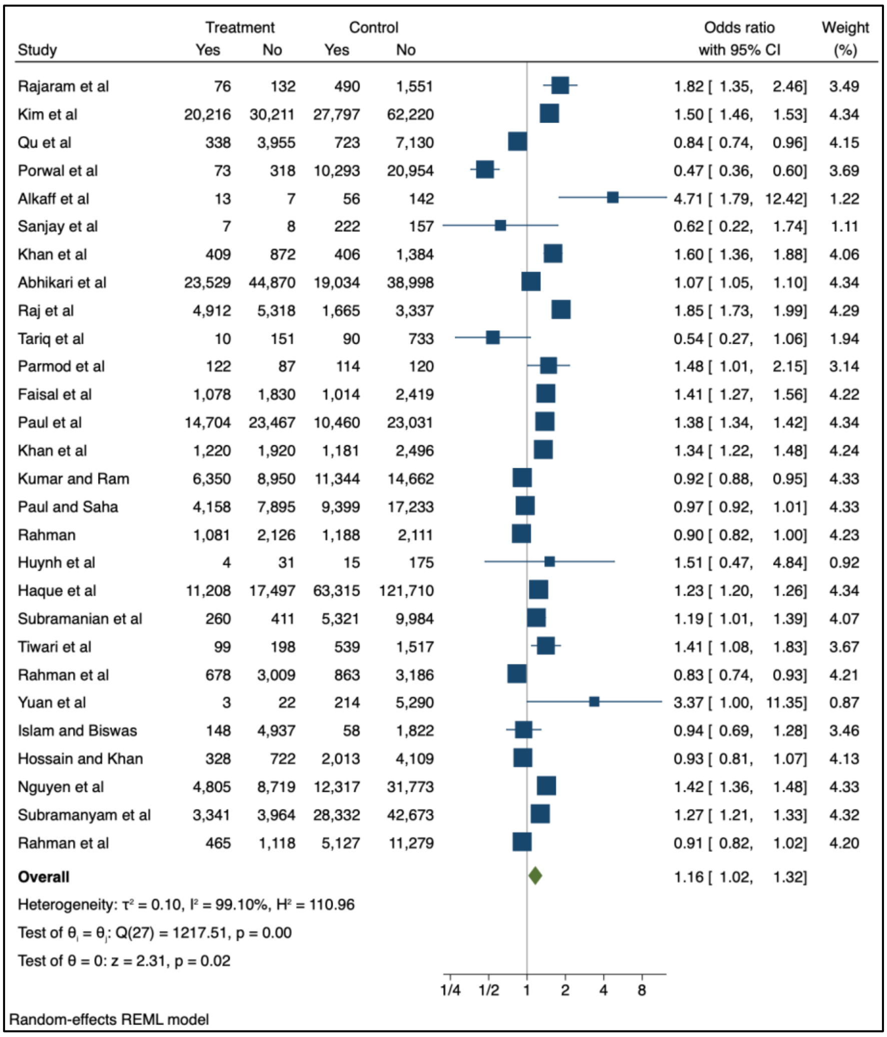


Key: Treatment – number of children in adolescent mother group, control – number of children in adult mother group, yes – number of children with the outcome (moderate underweight, weight-for-age z-score (WAZ) <-2 to -3), no – number of children without the outcome

**Supplementary Figure 6. Association between adolescent pregnancy (10-24 years) and the pooled odds of severe childhood wasting (1-59 months), versus adult pregnancy, stratifieted by region (Asia)**

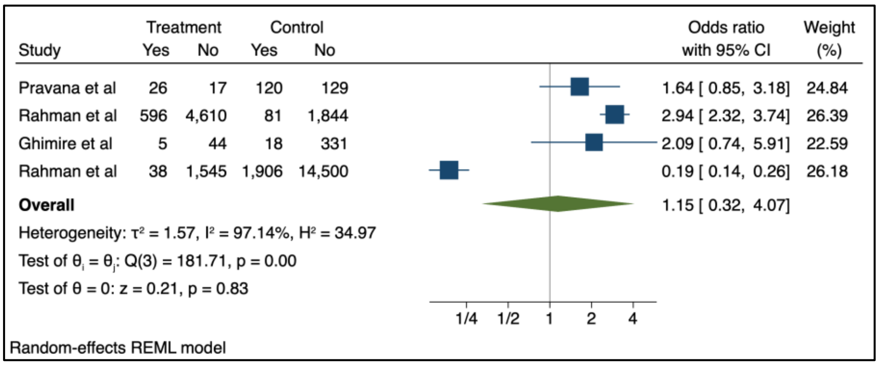


Key: Treatment – number of children in adolescent mother group, control – number of children in adult mother group, yes – number of children with the outcome (severe wasting, weight-for-height z-score (WHZ) or weight-for-length z-score (WLZ) <-3 or mid-upper arm circumference (MUAC) ≤115mm), no – number of children without the outcome

**Supplementary Figure 7. Association between adolescent pregnancy (10-24 years) and the pooled odds of severe childhood underweight (1-59 months), versus adult pregnancy, stratified by region (Asia)**

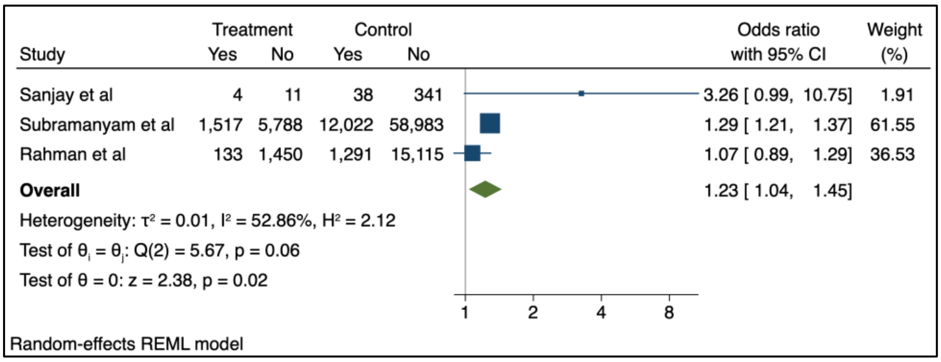


Key: Treatment – number of children in adolescent mother group, control – number of children in adult mother group, yes – number of children with the outcome (severe underweight, weight-for-age z-score (WAZ) <-3), no – number of children without the outcome

**Supplementary Figure 8. Association between adolescent pregnancy (10-24 years) and the pooled odds of moderate childhood wasting (1-59 months), versus adult pregnancy, stratified by region (sub-Saharan Africa)**

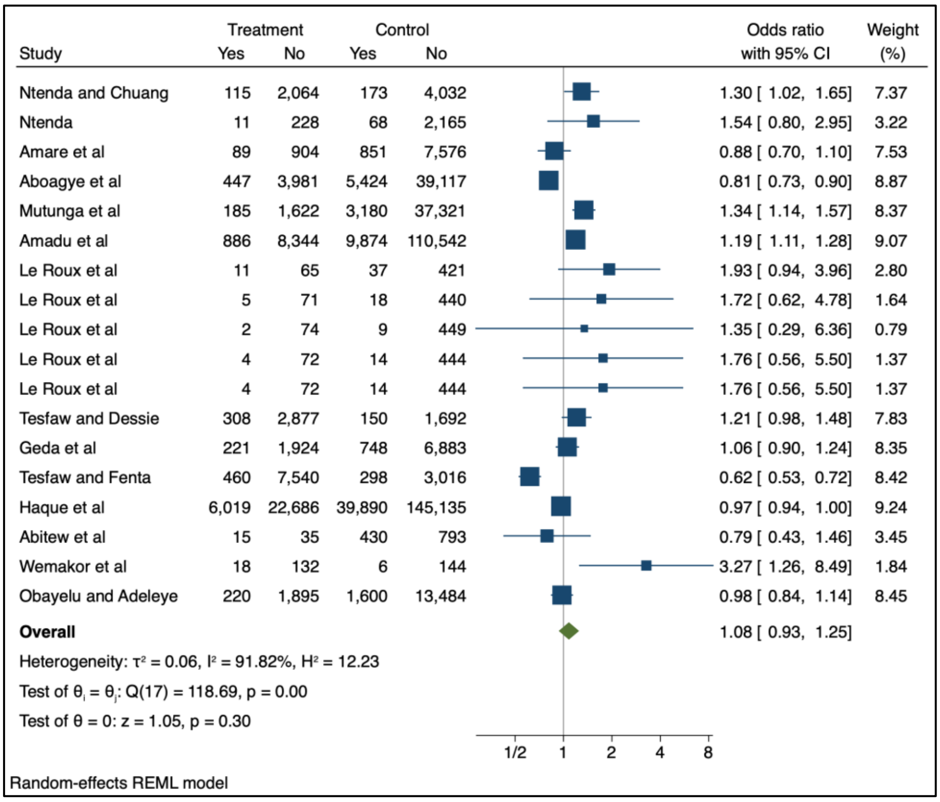


Key: Treatment – number of children in adolescent mother group, control – number of children in adult mother group, yes – number of children with the outcome (moderate wasting, weight-for-height z-score (WHZ) or weight-for-length z-score (WLZ) <-2 to -3 or mid-upper arm circumference (MUAC) 125mm to >115mm), no – number of children without the outcome

**Supplementary Figure 9. Association between adolescent pregnancy (10-24 years) and the pooled odds of moderate childhood underweight (1-59 months), versus adult pregnancy, stratified by region (sub-Saharan Africa)**

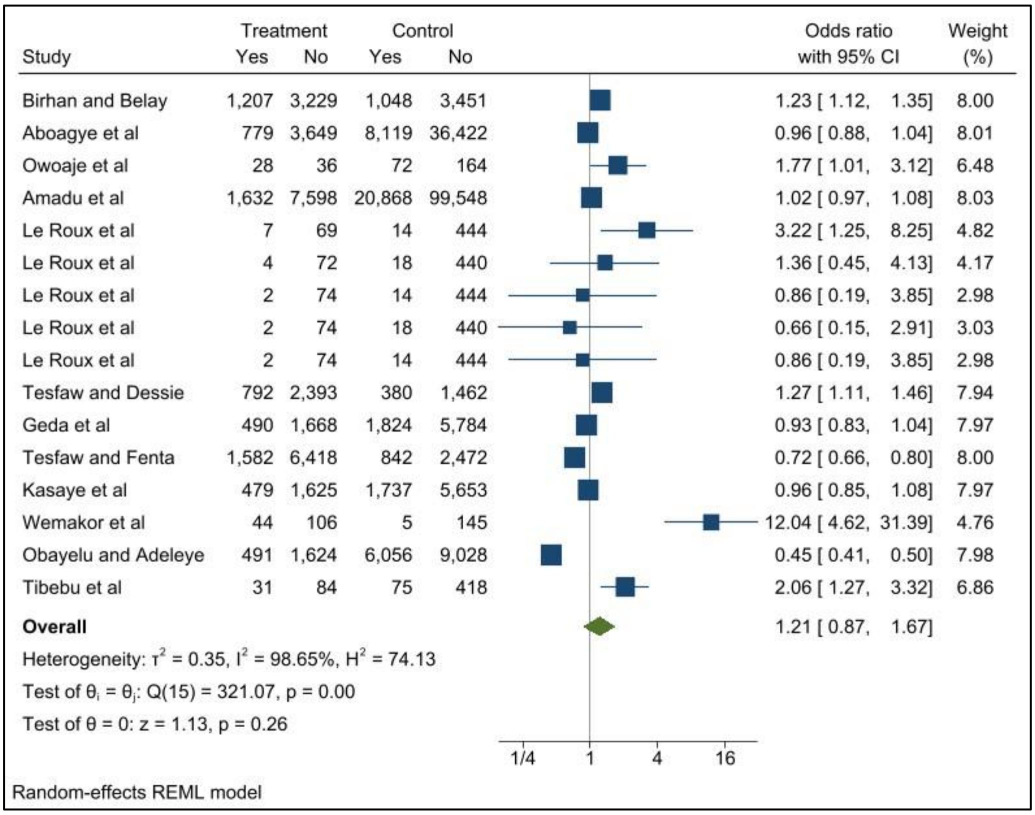


Key: Treatment – number of children in adolescent mother group, control – number of children in adult mother group, yes – number of children with the outcome (moderate underweight, weight-for-age z-score (WAZ) <-2 to -3), no – number of children without the outcome

**Supplementary Figure 10. Association between adolescent pregnancy (10-24 years) and the pooled odds of severe childhood wasting (1-59 months), versus adult pregnancy, stratified by region (sub-Saharan Africa)**

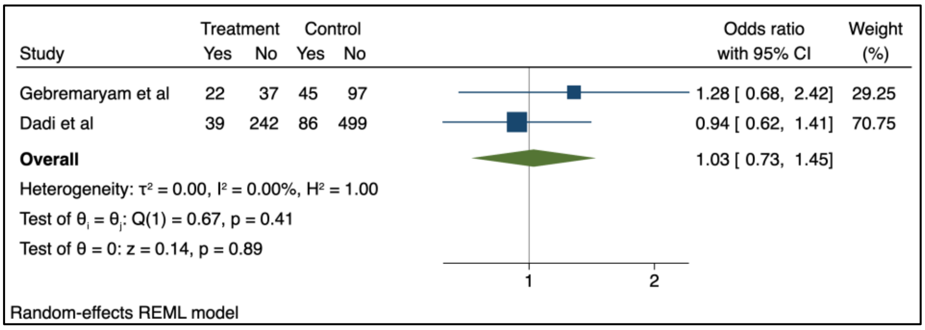


Key: Treatment – number of children in adolescent mother group, control – number of children in adult mother group, yes – number of children with the outcome (severe wasting, weight-for-height z-score (WHZ) or weight-for-length z-score (WLZ) <-3 or mid-upper arm circumference (MUAC) ≤115mm), no – number of children without the outcome

**Supplementary Appendix 5. Summary of findings from studies included in the qualitative synthesis**

**a. Moderate and severe wasting**

| Ref | First Author | Year | Study Design | Infant Age categories (months) | Country | Study Size | Maternal age category <20 years | Key findings | Indication of childhood wasting outcomes* |
|---|---|---|---|---|---|---|---|---|---|
| **Moderate wasting** | | | | | | | | | |
| 2 | Nguyen | 2021 | Cross-sectional | 0 - 59 | Bangladesh | 12,006 | ■ | Adjusted regression model for age at first birth:<br>10 – 15 years: -0.03 (-0.12, 0.06)<br>p = not significant<br>16 – 19 years: 0.01 (-0.05. 0.07)<br>p = not significant | ⇔ |
| 4 | Friebert | 2017 | RCT | Outcomes measured at 6 weeks & 12 weeks | Malawi | 2284 | ■ | Mean WLZ (SD)<br>At 6 weeks:<br>Mother <18 years: 0.3 (±1.1)<br>Mother 18 - 20 years: 0.4 (±1.0)<br>Mother >20 years: 0.3 (±1.1)<br>P value = 0.33<br><br>At 12 weeks:<br>Mother <18 years: 0.3 (±1.0)<br>Mother 18 - 20 years: 0.3 (±1.0)<br>Mother >20 years: 0.1 (±1.1)<br>p = 0.01<br>**NB:** all mothers were malnourished | ⇔ |
| 8 | Fuada | 2020 | Cross-sectional | 0 - 59 | Indonesia | 978 | ■ | OR for adolescent mothers ≤15 years compared to mothers aged 16-19 years: 1.810 (0.648 - 5.055) p = 0.402 | ⇑ |
| 10 | Issah | 2022 | Cross-sectional | 0 - 59 | Nigeria | 2425 | ⊕ | aOR<br>15 – 24 years: reference group<br>25 - 34: 0.66 (0.32 - 1.36) P = 0.260<br>35 - 49: 1.16 (0.38 - 3.56) p = 0.798 | ⇑ |
| 11 | Fall | 2015 | Cohort | Outcomes measured at 2 years | Brazil, Guatemala, India, the Philippines, | 19403 | ■ | aORs<br>≤19 years: 1.12 (0.86 – 1.46)<br>20 – 24 years: reference group<br>25 – 29 years: 0.91 (0.75 0 1.11)<br>30 – 34 years: 1.05 (0.84 – 1.31) | ⇑ |

| | | | | | | | | | |
|---|---|---|---|---|---|---|---|---|---|
| | | | | | and South<br>Africa | | | ≥35 years: 0.97 (0.74 – 1.28)<br>P = 0.64 | |
| 13 | Hiruy | 2021 | Cross-<br>sectional | 6 - 23 | Ethiopia | 8003 | ■ | The study shows the trend in aOR over the years:<br>15 – 19 years: Reference Group<br>**2000**<br>20 – 29 years: 1.03 (0.66-1.63)<br>30 – 39 years: 1.19 (0.74 – 1.90)<br>40 – 49 years: 1.06 (0.59 – 1.92)<br>**2005**<br>20 – 29 years: 0.97 (0.43 – 2.18)<br>30 – 39 years: 1.41 (0.61 – 3.25)<br>40 – 49 years: 1.79 (0.62 – 5.21)<br>**2011**<br>20 – 29 years: 1.02 (0.65 – 1.60)<br>30 – 39 years: 1.09 (0.69 – 1.75)<br>40 – 49 years: 1.36 (0.75 – 2.45)<br>**2016**<br>20 – 29 years: 0.75 (0.45 – 1.24)<br>30 – 39 years: 0.55* (0.34 – 0.90)<br>40 – 49 years: 0.84 (0.44 – 1.61)<br>*p = <0.05 | More recently:<br>⇑ |
| 14 | Ali | 2019 | Cross-<br>sectional | 0 - 59 | Bangladesh | 6468 | ⊕ | aOR<br>15 – 24 years: reference group<br>25 - 49: 1.06 p = 0.419 | ⇓ |
| 27 | Ahmed | 2012 | Cross-<br>sectional | 0 - 24 | Bangladesh | 8858 | ■ | aOR<br><20 years: 1.12 (0.81 – 1.57)<br>20 – 30 years: reference group<br>>30 years: 1.38 (0.96 – 1.99) | ⇑ |
| 30 | Sobkoviak | 2012 | Cross-<br>sectional | 0 - 48 | Liberia | 2467 | ⊕ | aOR<br>15 – 24 years: reference group<br>25 - 34: 1.45 (SE: 0.44)<br>35 - 49: 0.68 (SE: 0.34) | ⇓ |
| 32 | Masibo | 2020 | Cross-<br>sectional | 0 - 59 | Kenya | 7830 | ■ | uOR <-2SD<br>Overweight/obese mother – wasted child<br><20 years: 0.73 (0.11 – 4.87) p 0.749<br>20 – 30 years: 0.94 (0.29 – 3.03) p 0.918<br>30 – 40 years 1.49 (0.45 – 4.94) p 0.509<br>40+ years: reference group | ⇓ |

| | | | | | | | | | |
|---|---|---|---|---|---|---|---|---|---|
| 36 | Boah | 2019 | Cross-sectional | 0 - 59 | Ghana | 2636 | ■ | 15 - 19 years: reference group.<br>20 - 29: 1.34 (0.30 - 6.03)<br>30 - 39: 1.30 (0.27 - 6.38)<br>40 - 49:2.51 (0.38 - 16.61)<br>No significant p-values reported | ⇓ |
| 37 | Mena-Melendez | 2020 | Cross-sectional | 0 - 59 | Latin America: Bolivia, Colombia, Guatemala and Peru | 15827 | ■ | Adjusted correlation co-efficient for age at first birth: 1.009 (0.821 – 1.240) SD: 0.106 | ⇑ |
| 39 | Poda | 2017 | Cross-sectional | 0 - 59 | Burkina Faso | 6337 | ■ | Univariate OR<br><20 years: reference group.<br>20 - 24: 0.61 (0.42 - 1.39)<br>25 - 29: 0.55 (0.38 - 1.29)<br>30 - 34: 0.54 (0.37 - 1.29)<br>>=35: 0.50 (0.34 - 0.72)<br>P=<0.01<br>Multivariate OR<br><20 years: reference group.<br>20 – 24: 0.91 (0.61 – 1.36)<br>25 – 29: 0.88 (0.57 – 1.36)<br>30 – 34: 0.91 (0.56 – 1.46)<br>≥35: 0.88 (0.52 – 1.50) | ⇑ |
| 40 | Mashal | 2008 | Cross-sectional | 0 - 59 | Afghanistan | 2472 | ■ | uOR<br>Married ≥16: reference group<br>Married <16: 1.18 (0.85 – 1.63) | ⇑ |
| 45 | Samuel | 2022 | Cross-sectional | 6 - 11 | Ethiopia, | 2036 | ⊕ | uOR<br><25 years: 0.60 (0.42 – 0.85) p = 0.004<br>>25 years: reference group | ⇓ |
| 46 | Akseer | 2018 | Cross-sectional | 0 - 59 | Afghanistan | 22158 | ■ | Crude coefficient:<br>40 – 49 years: reference group<br>30 – 39 years: -0.08 (-0.22 – 0.05)<br>20 – 29 years: -0.12 (-0.25 – 0)<br>15 – 19 years: -0.07 (-0.26 – 0.12) | ⇑<br>(compared to ref. grp only) |
| 56 | Beiersmann | 2013 | Cross-sectional | 6 - 51 | Burkina Faso | 460 | ■ | Mean (SD) WLZ<br><20 years: -1.68 (1.03)<br>20 – 24 years: -1.20 (1.15) | ⇑ |

| | | | | | | | | | |
|---|---|---|---|---|---|---|---|---|---|
| | | | | | | | | 25 – 29 years: -1.04 (1.19)<br>30 – 34 years: -1.10 (1.25)<br>>35 years: -1.34 (1.28)<br>P 0.61 | |
| 57 | Ickes | 2015 | Cross-sectional | 0 - 23 | Uganda | 1897 | ■ | aOR<br>First birth >16 years: reference group<br>2006<br>First birth <16 years: 0.96 (0.62 – 1.49)<br>2011<br>First birth <16 years: 1.00 (0.57 – 1.75) | ⇔ |
| 63 | Nguyen | 2017 | Cross-sectional | 0 - 5 | Bangladesh | 2000 | ■ | Adjusted mean WHZ ±SD<br>≤19 years: -0.68 ±1.60<br>>19 yrs) = -0.75 ±1.39 | ⇔ |
| 64 | Hien and Hoa | 2009 | Cross-sectional | 6 - 36 | Vietnam | 383 | ⊕ | aORs:<br>>24 years: reference group<br>≤24 years: 2.16 (0.88 – 5.29) | ⇑ |
| 65 | Hien and Kam | 2008 | Cross-sectional | 0 - 59 | Vietnam | 650 | ⊕ | aORs:<br>25 – 34 years: reference group<br>≤24 years: 1.20 (0.65 - 2.21) | ⇑ |
| 67 | Olodu | 2019 | Cross-sectional | 6 - 59 | Nigeria | 300 | ■ | This study assessed the nutritional status of U5 children born to teenage mothers. 25.3% (72/285) of the children were wasted, but there is no comparison group provided. | N/A |
| 70 | Biswas | 2021 | Cross-sectional | 0 - 59 | South and South-East Asian countries | 798961 | ■ | aOR <-2SD<br>Overweight/obese mother & wasted child:<br><20 years: reference group<br>20 – 29 years: 2.56 (2.11 - 3.1) p <0.005<br>30 – 39 years: 3.67 (3.03 - 4.46) p <0.005<br>40 – 49 years: 3.90 (3.14 - 4.85) p <0.005 | ⇓ |
| 72 | Kasaye | 2019 | Cross-sectional | 0 – 59 | Ethiopia | 9494 | ⊕ | Proportions were available, however this is a duplicated data source, so removed from the meta-analysis.<br>Crude OR for mother's age:<br>15 – 24 years: 1.06 (0.90 – 1.24)<br>>24 years: reference group<br>P = 0.4872 | ⇑ |
| 78 | Das | 2019 | Cross-sectional | 0 - 59 | Bangladesh | 5951 | ■ | aOR <-2SD<br>Overweight/obese mother & wasted child: | ⇔ |

| | | | | | | | | | |
|---|---|---|---|---|---|---|---|---|---|
| | | | | | | | | ≤15 years: reference group<br>16 – 20 years: 1.17 (0.56 – 2.82)<br>21 – 25 years: 1.08 (0.39 – 3.08)<br>≥26 years: 0.93 (0.16 – 4.06) | |
| 80 | Mokwena | 2022 | Cross-sectional (hospital based study) | 1 - 36 | South Africa | 94 | ■ | All participants were malnourished.<br>Disaggregated by maternal age the proportions are:<br>16 – 20 years: 19%<br>21 – 25 years: 35%<br>26 – 30 years: 26%<br>31 – 40 years: 14%<br>NB: There are no 'healthy' controls included. | N/A |
| 81 | Gbadamosi | 2017 | Cross-sectional | 1 - 12 | South Africa | 186 | ■ | aOR:<br>≥19 years: reference group<br><19: 1.01 (0.232 - 4.403) | ⇔ |
| 86 | Bekele | 2021 | Cross-sectional | 0 - 59 | Ethiopia | 21514 | ⊕ | Absolute contribution (AC) and percentage contribution (PC) show the adjusted contributions to inequalities for each predictor, for both 2000 and 2016:<br>**2000**<br>15 – 24 years: reference group<br>25 – 34 years: AC <0.0001 PC -0.71<br>35 – 44 years: AC <0.0001 PC -0.17<br>45 – 49 years: AC <0.0001 PC -1.71<br>**2016**<br>15 – 24 years: reference group<br>25 – 34 years: AC <0.0001 PC -0.03<br>35 – 44 years: AC <0.0001 PC -0.17<br>45 – 49 years: AC -0.001 PC 2.05 | ⇔ |
| 88 | Gewa and Yandell | 2012 | Cross-sectional | 0 - 60 | Kenya | 1851 | ⊕ | Mean maternal age of mothers first birth for children that were wasted versus children that were not wasted.<br>**0 - 24-month-old children:**<br>Wasted: (n = 147) 19 (18.9 - 19.2)<br>not wasted: (n = 1704) 18.2 (17.7 - 18.7)<br>**25 - 60-month-old children:**<br>Wasted: (n = 95), 19.1 (18.3 - 19.9)<br>not wasted: (n=1846) 19.1 (19.0 - 19.3) | N/A |

| | | | | | | | | | |
|---|---|---|---|---|---|---|---|---|---|
| **Severe wasting** | | | | | | | | | |
| 27 | Ahmed | 2012 | Cross-sectional | 0 - 24 | Bangladesh | 8858 | ■ | aOR<br><20 years: 0.96 (0.08 – 1.15)<br>20 – 30 years: reference group<br>>30 years: 0.96 (0.77 – 1.19) | ⇔ |
| 80 | Mokwena | 2022 | Cross-sectional (hospital based study) | 1 - 36 months | South Africa | 94 | ■ | All participants were malnourished.<br>Proportion of those admitted with severe wasting in each maternal age group versus moderate wasting<br>16 – 20 years: 44.4%<br>21 – 25 years: 32.3%<br>26 – 30 years: 23.1%<br>31 – 40 years: 50% (NB n=2)<br>NB: There are no 'healthy' controls included. | N/A |

■ - Yes ⊕ - No
aOR: adjusted odds ratio, uOR: unadjusted odds ratio
p: p-value
*Indication of outcome direction:
⇑ = study indicates adolescent pregnancy is associated with **increased** childhood wasting, compared to adult pregnancy
⇔ = study reports no diifference between the groups
⇓ = study indicates adolescent pregnancy is associated with **reduced** childhood wasting compared to adult pregnancy

### b. Moderate and severe underweight

| Ref | First Author | Year | Study Design | Infant Age categories (months) | Country | Study Size | Maternal age category <20 years | Key findings | Indication of childhood underweight outcomes* |
|---|---|---|---|---|---|---|---|---|---|
| **Moderate underweight** | | | | | | | | | |
| 3 | Schott | 2017 | Cohort | 0 - 78 | Ethiopia, India | 253 | ■ | This study was an intergenerational study that assessed the association between adolescent mothers' anthropometric measurements at aged 8, 12, 15 and 19 years, as well as maternal diet at the age of 15 years (and grandmother's characteristics) on their off-springs WAZ (HAZ and birthweight).<br>**Mother's age at birth predicted higher cWAZ between birth and 2013 ($\beta$0.46 p<0.10 and $\beta$ 0.53 p<0.05)**<br>There was no comparison with an adult cohort. | ⇑* |
| 4 | Friebert | 2017 | RCT (assessor blinded) | 6 weeks and 12 weeks | Malawi | 2284 | ■ | Mean WAZ (SD)<br>At 6 weeks:<br>Mother <18 years: -1.1 +-1.2<br>Mother 18 - 20 years: -0.9 +-1.1<br>Mother >20 years: 0.8 +-1.0<br>P = <0.00<br><br>At 12 weeks:<br>Mother <18 years: -0.8 +- 1.1<br>Mother 18 - 20 years: -0.8 +-1.0<br>Mother >20 years: -1.0 +-1.1<br>p <0.00<br><br>NB: All mothers were malnourished | ⇑*<br>(At 6 weeks) |
| 8 | Fuada | 2020 | Cross-sectional | 0 - 59 | Indonesia | 978 | ■ | OR for Adolescent mother group<br>≤15 years: 1.311 (0.498 - 3.450)<br>15 – 19 years: reference group<br>p 0.773<br><br>**NB** – no adult mother comparison | ⇑ |

| | | | | | | | | | |
|---|---|---|---|---|---|---|---|---|---|
| 13 | Hiruy | 2021 | Cross-sectional | 6 - 23 | Ethiopia | 8003 | ■ | aOR provided for 2000, 2005, 2011, 2016.<br>15 – 19 years: reference group<br><br>2000<br>20 – 29 years: 1.45 (0.98 – 2.14)<br>30 – 39 years: 1.79 (1.20 – 2.68)**<br>40 – 49 years: 1.75 (1.06 – 2.88)*<br><br>2005<br>20 – 29 years: 1.19 (0.59 – 2.38)<br>30 – 39 years: 1.32 (0.64 – 2.73)<br>40 – 49 years: 2.26 (0.91 – 5.61)<br><br>2011<br>20 – 29 years: 1.18 (0.81 – 1.73)<br>30 – 39 years: 1.28 (0.86 – 1.91)<br>40 – 49 years: 1.28 (0.76 – 2.15)<br><br>2016<br>20 – 29 years: 0.54 (0.33 – 0.89)*<br>30 – 39 years: 0.67 (0.42 – 1.07)<br>40 – 49 years: 1.17 (0.65 – 2.21)<br><br>*p <0.05<br>**p <0.01 | ⇑* |
| 14 | Ali | 2019 | Cross-sectional | 0 - 59 | Bangladesh | 6468 | ⨁ | aOR:<br>15 – 24 years: reference group<br>25 - 49 years: 1.08<br>p 0.186 | ⇓ |
| 18 | Kumar and Paswan | 2021 | Cross-sectional | 0 - 59 | India | 11858 | ■ | aOR provided for 2005-2006 and 2015 -2016<br><20 years: reference group<br><br>**2005 - 2005**<br>20 – 29 years: 0.93 (0.523 – 1.654)<br>30 – 49 years: 0.811 (0.443 – 1.485)<br><br>**2015 - 2016**<br>20 – 29 years: 1.021 (0.923 – 1.130)<br>30 – 49 years: 1.021 (0.916 – 1.139) | ⇔ |

| | | | | | | | | | |
|---|---|---|---|---|---|---|---|---|---|
| 21 | Linnemayr | 2008 | Cross-sectional | 0 - 35 | Senegal | 4296 | ⊕ | OLS (ordinary least squares) regression model (Standard error) for ‘teenage mother’<br>Adjusted for individual and household variables:<br>-0.135** (0.053)<br>Adjusted for individual, household and community variables:<br>-0.122** (0.053)<br>Adjusted for ‘instrumented variables’ (the presence of either an NGO or health post in the village):<br>-0.113** (0.055)<br><br>**significance at 5% level | ⇑* |
| 27 | Ahmed | 2012 | Cross-sectional | 0 - 24 | Bangladesh | 8858 | ■ | aORs <-2SD:<br><20 years: 1.04 (0.86 - 1.26)<br>20 – 30 years: reference group<br>>30 years: 1.31 (1.04 - 1.65)*<br>*p <0.05 | ⇑ |
| 30 | Sobkoviak | 2012 | Cross-sectional | 0 - 48 | Liberia | 2467 | ⊕ | aOR:<br>15 – 24 years: reference group:<br>25 - 34: 0.65 (SE: 0.11) p <0.001<br>35 - 49: 0.41 (SE: 0.12) p <0.001 | ⇑* |
| 31 | Hossain | 2020 | Cross-sectional | 24 - 59 | 5 South Asian countries (India Bangladesh, Pakistan, Maldives and Nepal) | 146996 | ■ | aOR:<br>20 - 24 years: reference group<br>**<20 years:**<br>Bangladesh: 1.1 (0.9 - 1.3)<br>India: 1.1 (1.0 - 1.1)<br>Maldives: 1.1 (0.8 - 1.5)<br>Nepal: 0.9 (0.7 - 1.2)<br>Pakistan: 1.0 (0.8 - 1.2)<br>**≥25 years:**<br>Bangladesh: 0.8 (0.5 – 1.1)<br>India: 0.8 (0.8 – 0.8)<br>Maldives: 0.8 (0.5 - 1.3)<br>Nepal: 0.8 (0.5 – 1.4)<br>Pakistan: 0.9 (0.7 – 1.3) | ⇑ |
| 36 | Boah | 2019 | Cross-sectional | 0 - 59 | Ghana | 2636 | ■ | aOR:<br>15 - 19 years: reference group<br>20 – 29 years: 0.50 (0.17 - 1.42) | ⇑ |

| | | | | | | | | | |
|---|---|---|---|---|---|---|---|---|---|
| | | | | | | | | 30 – 39 years: 0.54 (0.16 - 1.76)<br>40 – 49 years: 0.76 (0.19 - 3.03) | |
| 32 | Masibo | 2020 | Cross-sectional | 0 - 59 | Kenya | 7830 | ■ | aOR<br>Overweight/obese mother – underweight child (<-2SD)<br><20 years: 0.43 (0.07 – 2.65) p 0.36<br>20 – 30 years: 0.55 (0.24 – 1.26) p 0.156<br>30 – 40 years: 1.08 (0.48 – 2.40) p 0.857<br>40+ years: reference group | ⇓ |
| 39 | Poda | 2017 | Cross-sectional | 0 - 59 | Burkina Faso | 6337 | ■ | Univariate and multivariate analysis:<br><20 years: reference group<br><br>**Univariate**<br>20 – 24 years: 0.68 (0.49 - 1.25)<br>25 – 29 years: 0.57 (0.42 - 1.01)<br>30 – 34 years: 0.61 (0.45 - 0.83)*<br>≥35 years: 0.61 (0.45 - 0.84)*<br><br>**Multivariate**<br>20 – 24 years: 0.77 (0.53 – 1.12)<br>25 – 29 years: 0.61 (0.41 – 1.18)<br>30 – 34 years: 0.61 (0.40 – 1.05)<br>≥35 years: 0.61 (0.38 – 0.97)*<br><br>*p <0.05 | ⇑ |
| 40 | Mashal | 2008 | Cross-sectional | 0 - 59 | Afghanistan | 2472 | ■ | uOR<br>Married >16 years: reference group<br>Married <16 years: 1.41 (1.12 - 1.78) | ⇑* |
| 46 | Akseer | 2018 | Cross-sectional | 0 - 59 | Afghanistan | 14000 | ■ | Mother's age at delivery was disaggregated by:<br><20 years<br>20 – 29 years<br>30 – 39 years<br>40 – 49 years<br>Using 30 – 39 years as the reference group it was found that mother's age was not significantly associated with underweight amongst the offspring. (non-significant ORs were not published) | ⇔ |

| | | | | | | | | | |
|---|---|---|---|---|---|---|---|---|---|
| 57 | Ickes et al | 2015 | Cross-sectional | 0 - 23 | Uganda | 1009 | ■ | ORs for 2006 and 2011:<br>First birth at >16 years old: reference group<br><br>**2006**<br>First birth <16 years: 1.03 (0.72 - 1.47)<br>**2011**<br>First birth <16 years 1.39 (0.92 - 2.10) | ⇑ |
| 63 | Nguyen | 2017 | Cross-sectional | 0 - 5 | Bangladesh | 200 | ■ | Adjusted mean WAZ ± SD:<br>≤19 years: -1.21 ±1.10<br>>19 years: -1.08 ±1.80 p <0.05 | ⇑ |
| 64 | Hien and Hoa | 2009 | Cross-sectional | 6 - 36 | Vietnam | 383 | ⊕ | aORs:<br>≤24 years: 1.12 (0.64 – 1.98)<br>>24 years: reference group | ⇑ |
| 65 | Hien and Kam | 2008 | Cross-sectional | 0 - 59 | Vietnam | 650 | ⊕ | aORs:<br>≤24 years: 0.93 (0.61 - 1.44)<br>25 – 34 years: reference group | ⇔ |
| 67 | Olodu | 2019 | Cross-sectional | 6 - 59 | Nigeria | 300 | ■ | This study assessed the nutritional status of U5 children born to teenage mothers. 29.5% (84/285) of the children were underweight, but there is no comparison group provided. | N/A |
| 68 | Nakamori | 2010 | Cross-sectional | 6 - 18 | Vietnam | 188 | ⊕ | ORs:<br><25 years: reference group<br>>25 years: 1.37 (0.60 - 3.14)<br>p 0.451 | ⇓ |
| 70 | Biswas | 2021 | Cross-sectional | 0 - 59 | South and South-East Asian countries | 798961 | ■ | Overweight/obese mother & underweight child:<br><20 years: reference group<br>20 – 29 years: 2.56 (2.11- 3.09) p <0.005<br>30 – 39 years: 3.65 (3.01 - 4.43) p <0.005<br>40 – 49 years: 3.87 (3.12 - 4.80) p <0.005 | ⇓ |
| 72 | Kasaye | 2019 | Cross-sectional | 0 – 59 | Ethiopia | 9494 | ⊕ | Proportions were available, however this is a duplicated data source, so removed from the meta-analysis.<br>Crude OR for mother's age:<br>15 – 24 years: 0.96 (0.85 – 1.08)<br>>24 years: reference group<br>P = 0.4798 | ⇓ |
| 78 | Das | 2019 | Cross-sectional | 0 - 59 | Bangladesh | 5951 | ■ | Overweight/obese mother & underweight child<br>≤15 years: reference group | ⇓ |

| | | | | | | | | | |
|---|---|---|---|---|---|---|---|---|---|
| 81 | Gbadamosi | 2017 | Cross-sectional | 1 - 12 | South Africa | 186 | ■ | 16 – 20 years: 1.47 (0.83 - 2.84)<br>21 – 25 years: 2.16 (1.08 - 4.48) p <0.05<br>≥26 years: 2.17 (0.72 - 5.73)<br>Not statistically significant<br>aOR:<br><19 years:1.25 (0.062 - 25.271)<br>≥19 years: reference group | ⇑ |
| 86 | Bekele | 2021 | Cross-sectional | 0 - 59 | Ethiopia | 21514 | ⊕ | Absolute contribution (AC) and percentage contribution (PC) show the adjusted contributions to inequalities for each predictor, for both 2000 and 2016:<br>**2000**<br>15 – 24 years: reference group<br>25 – 34 years: AC <0.0001 PC -0.10<br>35 – 44 years: AC <0.0001 PC -0.22<br>45 – 49 years: AC -0.011 PC 12.38<br>**2016**<br>15 – 24 years: reference group<br>25 – 34 years: AC <0.0001 PC -0.01<br>35 – 44 years: AC <0.0001 PC -0.03<br>45 – 49 years: AC -0.001 PC 0.77 | ⇔ |
| 88 | Gewa and Yandell | 2012 | Cross-sectional | 0 - 60 | Kenya | 1851 | ⊕ | Mean maternal age of mothers first birth for children that were wasted versus children that were not wasted.<br>**0 - 24-month-old children:**<br>Underweight: (n = 230) 18.2 (17.7 0 18.6)<br>not underweight: (n = 1621) 19.1 (18.9 – 19.3)<br>**25 - 60-month-old children:**<br>Underweight: (n = 338), 18.5 (18.1 – 18.8)<br>not wasted: (n=1604) 19.3 (19.1 – 19.4) | N/A |
| **Severe underweight** | | | | | | | | | |
| 27 | Ahmed et al | 2012 | Cross-sectional | 0 – 24 | Bangladesh | 8858 | ■ | aORs <-3SD:<br><20 years: 1.11 (0.95 – 1.29)<br>20 – 30 years: reference group<br>>30 years: 1.21 (1.02 – 1.43) | ⇑ |

■ - Yes ⊕ - No
aOR: adjusted odds ratio, uOR: unadjusted odds ratio
p: p-value

*significance of <0.05
*Indication of outcome direction:
⇑ = study indicates adolescent pregnancy is associated with **increased** childhood underweight, compared to adult pregnancy
⇔ = study reports no diifference between the groups
⇓ = study indicates adolescent pregnancy is associated with **reduced** childhood underweight compared to adult pregnancy
